# International Travel-Related Control Measures to contain The Covid-19 Pandemic: An update to a Cochrane Rapid Review

**DOI:** 10.1101/2022.03.24.22271703

**Authors:** Ameer Hohlfeld, Leila Abdullahi, Ahmed M. Abou-Setta, Mark E Engel

## Abstract

**Background:** COVID-19 has proven to be more difficult to manage for many reasons including its high infectivity rate. One of the potential ways to limit its spread is by controlling international travel. The objective of this systematic review is to identify, critically-appraise and summarize evidence on international travel-related control measures.

**Methods:** This review is based on the Cochrane review: *International travel-related control measures to contain the COVID-19 pandemic* and followed the same methods. In brief, we searched for clinical and modelling studies in general health and COVID-19-specific bibliographic databases. The primary outcome categories were (i) cases avoided, (ii) a shift in epidemic development and, (iii) cases detected. Secondary outcomes were other infectious disease transmission outcomes, healthcare utilisation, resource requirements and adverse effects if identified in studies assessing at least one primary outcome.

**Results:** We assessed 66 full-text articles that met with our inclusion criteria. Seventeen new studies (modelling = 9, observational = 8) were identified in the updated search. Most studies were of critical to moderate risk of bias. The added studies did not change the main conclusions of the Cochrane review nor the quality of the evidence (very low to low certainty). However, it did add to the evidence base for most outcomes.

**Conclusions:** Weak evidence supports the use of international travel-related control measures to limit the spread of COVID-19 via air travel. Real-world studies are required to support these conclusions.

## Background

In May 2020, WHO Guideline Development Group (GDG) identified the effectiveness of quarantine, microbiological detection, and quarantine with microbiological detections as potential interventions to mitigate transmission of SARS-CoV-2 as a key public health intervention in need of immediate guidance. In 2020 Burns et al published the first of two Cochrane Rapid Reviews on this topic (1). During 2020, the body of published and unpublished literature on COVID-19-related interventions grew exponentially prior to the international vaccine rollout. Subsequently, an updated review was published at the start of 2021. With the continuous growth in research, we now present an update to the 2021 Cochrane review. The objective of this systematic review is to identify, critically-appraise and summarize evidence on border closures and travel restrictions to control the spread of SARS-CoV-2 transmission between countries and regions.

## Methods

The research question was defined using the PICO format. We employed similar methods as per Burns 2021 (1).

### Criteria for considering studies for this review

We used the same criteria set out by Burns 2021 (1). Studies comprising:

Population: Human populations (without any age restriction) susceptible to SARS-CoV- 2/COVID-19.

Intervention: Travel-related control measures (specifically quarantine, microbiological detection or a combination thereof) affecting human travel across national borders.

Outcome(s): ***Cases avoided*** due to the measure (e.g., number, proportion, rate of cases observed or predicted in the community with and without the intervention).

***Shift in epidemic development*** due to the intervention (e.g., probability of epidemic, time to/delay in epidemic arrival or peak, size of epidemic peak, change in the effective reproduction number).

***Cases detected*** due to the measure: we focused on outcomes we felt are most relevant for decision-makers in the current pandemic: the proportion of cases detected among the total number of cases (i.e., sensitivity, case detection rate) and the proportion of cases among those screening positive (i.e., the positive predictive value).

Types of studies: randomized trials, non-randomized trials, observational studies, and modelling studies on the effects of travel-related control measures affecting human travel during the COVID-19 pandemic

### Search methods for identification of studies

We adapted the search strategies from Burns 2021, by excluding terms not-related to COVID- 19 (such as MERS, H1N1, SARS01). Searches were conducted from the start of November 2020 until November 2021. We did not add language restrictions to our search. We searched Scopus, Pubmed, the Cochrane COVID-19 WHO COVID-19 Global Literature on Coronavirus disease, MedRXiv and BioRXiv. Due to our limited available databases, we could not search Ovid MEDLINE and Epub Ahead of Print, In-Process & Other Non-Indexed Citations, Daily and Versions, Ovid Embase.

## Data collection and analysis

### Selection of studies

Two reviewers used the Rayyan software to screen titles and abstracts independently and in duplicate (2). Conflicting decisions were moved to full text eligibility assessment. Discrepancies were discussed among the review team. Data extraction, risk of bias in epidemiological studies and assessment of the quality of modelling studies was divided amongst the review team and cross-checked by at least one other reviewer for correctness and consistency. The two reviewers involved in these procedures discussed any discrepancies. The review team recorded the reasons for excluding full-text studies at the final selection stage. It was essential that the author team reached consensus at every stage of this review. Thus, reviewers ensured that the highest methodological rigour was maintained. Each stage of the review was piloted. To ensure reporting accuracy, the team regularly met to discuss queries or difficulties arising during the review process.

### Data extraction and management

Data extraction forms where based on those from Burns 2021 (1). We designed these using Microsoft Excel. We piloted the data extraction forms and recorded data from included studies. All review authors extracted study characteristics independently but not in duplicate. Reviewers checked each other’s data extraction for accuracy.

### Assessment of risk of bias in included studies

We followed the same process of assessing risk of bias as set out by Burns 2021 (1). Briefly, one reviewer rated the risk of bias, a second reviewer checked the judgements. We assessed risk of bias for observational studies using ROBINS-I (3). There are no validated tools available to assess risk of bias of modelling studies; we used the bespoke assessment tool described previously (1, 4).

### Data synthesis

It was predicted that we would find observational studies and modelling studies. We separated these two studies designs and considered these as independent bodies of evidence (1). Furthermore, regardless of study design, it was expected that we would find extensive heterogeneity across all studies’ setting, population, intervention, and other contextual factors, as well as study methods. This meant that data would not be sufficiently similar to conduct meta-analyses. Therefore, we planned to descriptively synthesise the data in tables. We stratified studies by study design which were then further stratified according to interventions and outcomes. Further details on data synthesis that we followed are detailed in Burns 2021(1).

### Assessment of heterogeneity and subgroup analyses

Consistent with Burns 2021, a meta-analysis was not undertaken. The substantial heterogeneity of included studies would not allow for subgroup analysis. Nevertheless, we planned to ascertain and report the potential sources of heterogeneity that may influence intervention effectiveness. Further details can be found in Burns 2021(1).

### Assessment of certainty of evidence

The certainty of evidence was rated using the GRADE approach (Grading of Recommendation, Assessment, Development and Evaluation) as high, moderate, low or very low (5). Two reviewers independently assessed the evidence for the primary outcomes followed by discussion with a second reviewer. Together they rated the certainty of evidence. We used the definitions described in Burns 2021 to guide our reasons and decisions for rating observational studies and modelling studies (1).

## Results

### Results of the search

We used the PRISMA flow diagram shown in Figure 1 to depict the study selection process (6). We screened 4423 unique titles and abstracts of which 66 full-text articles were assessed for eligibility. Seventeen studies met the eligibility criteria. We descriptively synthesised these in the Summary of Findings (SoF) – Tables 1-3. Characteristics of included studies are provided in Table 4. We excluded 49 studies, reasons for exclusion are provided in Table 5.

**Figure 1.**
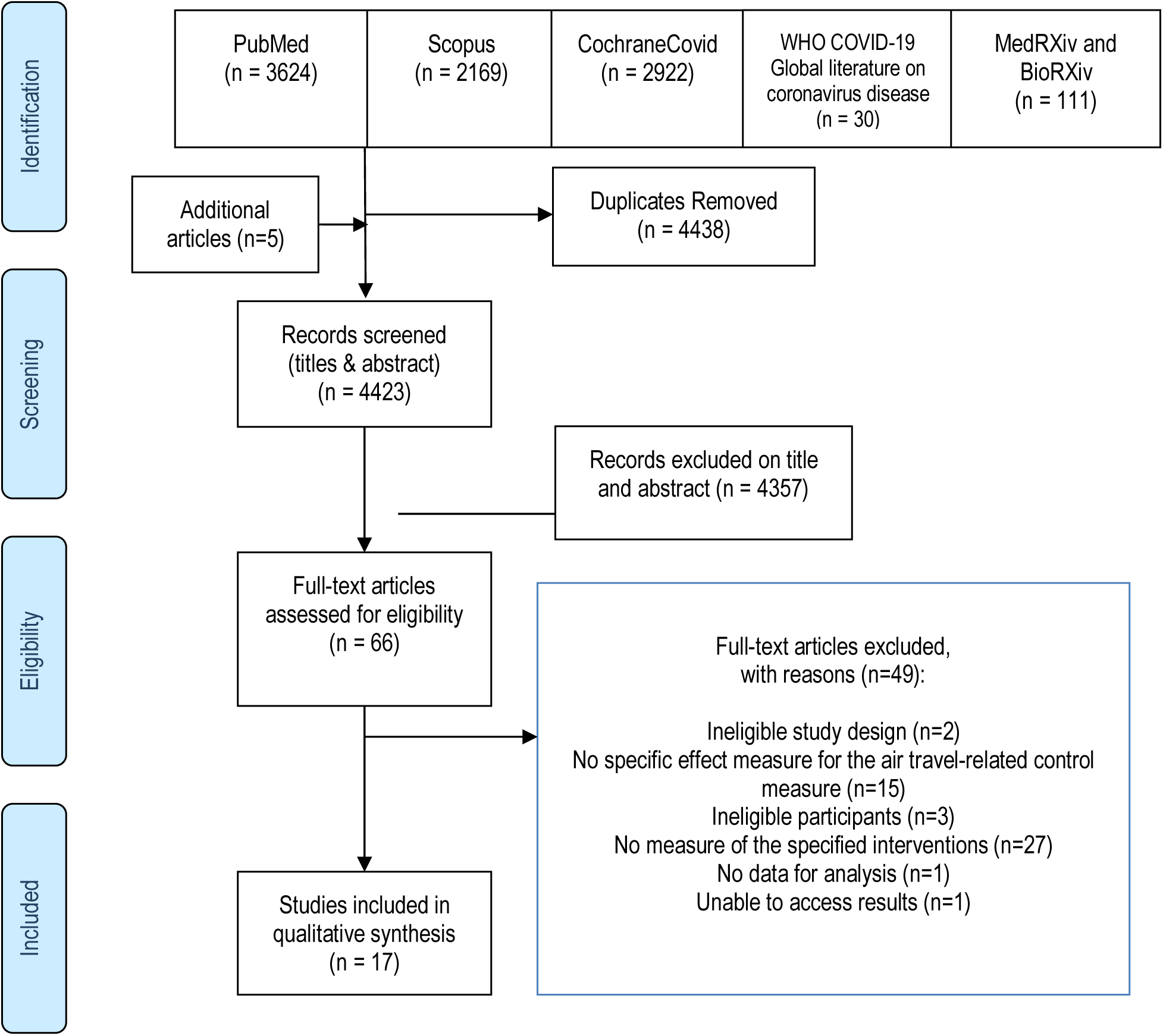
Schematic PRISMA flow diagram of the literature search.

**Table 1:**
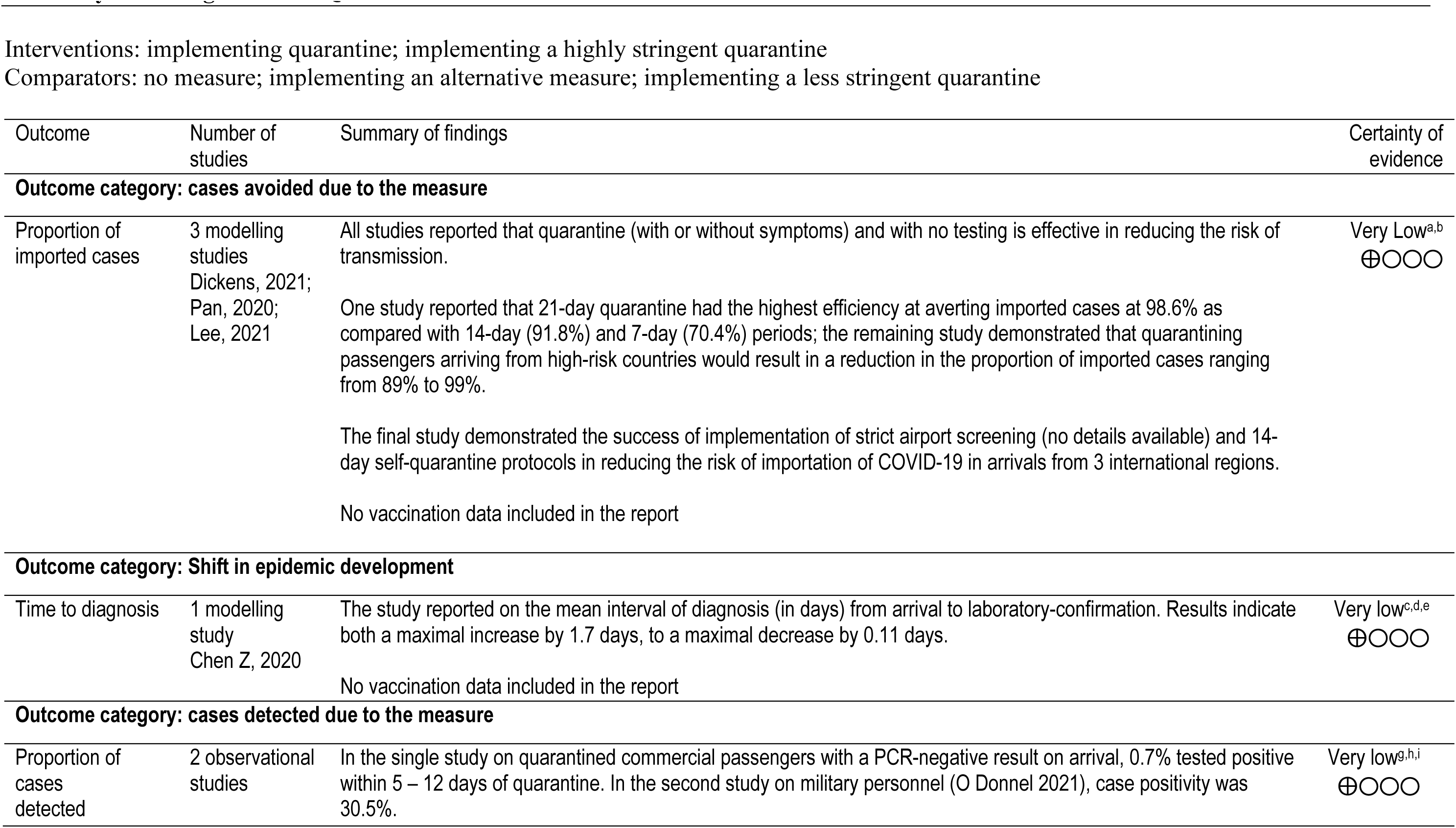
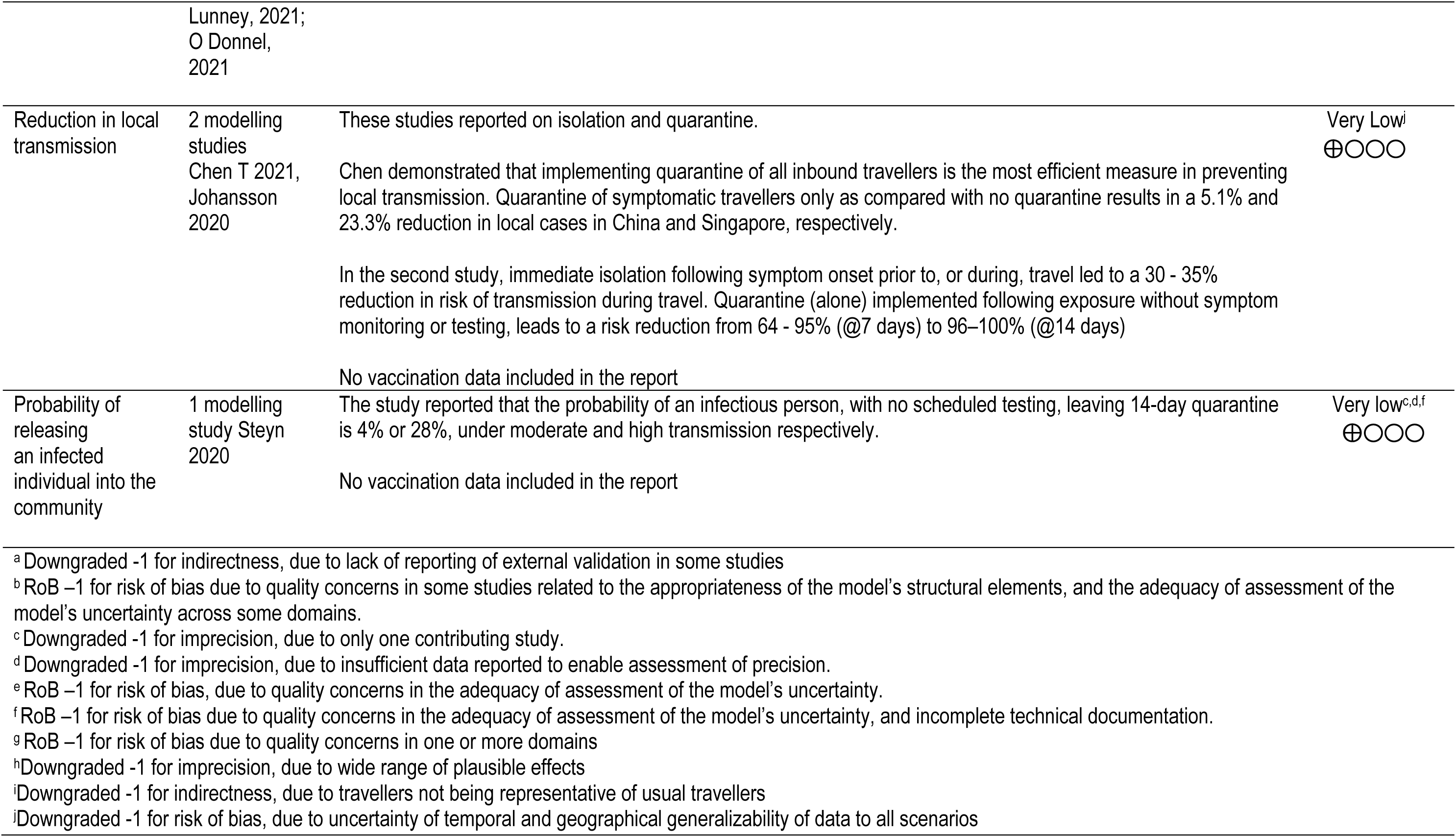
Quarantine

**Table 2:** Microbiological Detection

| Outcome | Number of studies | Summary of findings | Certainty of evidence |
| --- | --- | --- | --- |
| **Outcome category: cases avoided due to the measure** |  |  |  |
| Number or proportion imported | 2 modelling studies<br>Dickens 2021, Steyn 2020 | Assuming that exposure may have occurred at any time in the 7 days prior to departure, pre-departure testing can reduce risk of transmission during travel from 10%–72%, with reduction increasing as the interval between testing and day of travel decreases.<br>The addition of entry testing to pre-departure testing improves the detection of cases imported from 57.2% (2-day pre-departure testing) to 84.8% (83.1% for rapid antigen pre-departure and on arrival). Results vary according to timing of pre-departure test.<br>One study showed that testing within a 1-day interval between pre-departure and arrival may give false negatives, with only 50% of infected travellers being detected.<br>No vaccination data included in the report | Very low<sup>a,b,c</sup><br>⊕○○○ |
| **Outcome category: Shift in epidemic development** |  |  |  |
| Median time to outbreak | 1 modelling study<br>Wilson 2021 | The study reported that pre-flight saliva testing (assuming a 62% sensitivity) will delay an outbreak by 0.3 years delay [(95% range of simulations: 4 days to 1.7 years); mean number of flights to outbreak = 170].<br>Increasing the number of testing days after arrival reduces the median waiting time to the next outbreak. For example, pre-flight testing plus PCR tests on days 1, 3 and 12 delayed an outbreak by 0.7y [(9d – 3.6y); mean number of flights to outbreak= 359] as compared with preflight testing and testing on day 1 only, 0.4y [(6d – 2.3y); 225 flights]. | Very Low<sup>d</sup><br>⊕○○○ |
|  |  | The best result is observed with pre-flight testing plus PCR tests on days 1, 3 and 12 combined with mask usage until the last test, symptom reporting and contact tracing resulting in a delay of 1.5 years [(20 days to 8.1 y); 802 flights] |  |
|  |  | No vaccination data included in the report |  |
| Risk of outbreak | 1 modelling study<br>Wilson 2021 | The study reported the annual risk of an outbreak with pre-flight saliva testing alone is 88.4%. Pre-flight testing on days 1, 3 and 12 combined with mask usage until the last test, symptom reporting, and contact tracing further reduced the risk of an outbreak to 36.6% | Very Low<sup>d</sup><br>⊕○○○ |
|  |  | No vaccination data included in the report |  |
| **Outcome category: cases detected due to the measure** |  |  |  |
| Proportion of cases detected | 5 observational studies<br>Cao-Lormeau 2021, Lunney 2021, Ohlsen 2020, Tande 2021, Zhang 2021 | Across the studies, the proportion of cases amongst all travellers detected ranged from 0.01% to 1.5% over the periods of the study. One of the studies reported success at utilizing self-collection of samples conducted at 4 days post-arrival. | Very Low<sup>e,f</sup><br>⊕○○○ |
|  |  | No vaccination data included in the report |  |
| Reduction in transmission | 1 modelling study<br>Johanssen 2020 | Pre-departure testing can reduce risk of transmission <u>during</u> travel between 10 - 72% depending on time interval between testing and day of travel. Variation in results is dependent on time of specimen collection, (with greatest effectiveness if taken on day of travel), and degree of sensitivity of the test, (with higher sensitivity test giving a higher reduction in risk). A single test reduces the risk of transmission <u>after</u> travel from 29-53%, regardless if performed at one or two days post-arrival. | Very Low<sup>d</sup><br>⊕○○○ |
|  |  | No vaccination data included in the report |  |
| <sup>a</sup> Downgraded -1 for indirectness, due to lack of reporting of external validation in some studies<br><sup>b</sup> Downgraded -1 for imprecision, due to insufficient data reported to enable assessment of precision.<br><sup>c</sup> RoB -1 for risk of bias due to quality concerns in the adequacy of assessment of the model's uncertainty, and incomplete technical documentation.<br><sup>d</sup> Downgraded -1 for imprecision, due to only one contributing study.<br><sup>e</sup> RoB -1 for risk of bias due to quality concerns in one or more domains<br><sup>f</sup> Downgraded -1 for indirectness, due to variation in testing methods |  |  |  |

**Table 3:**
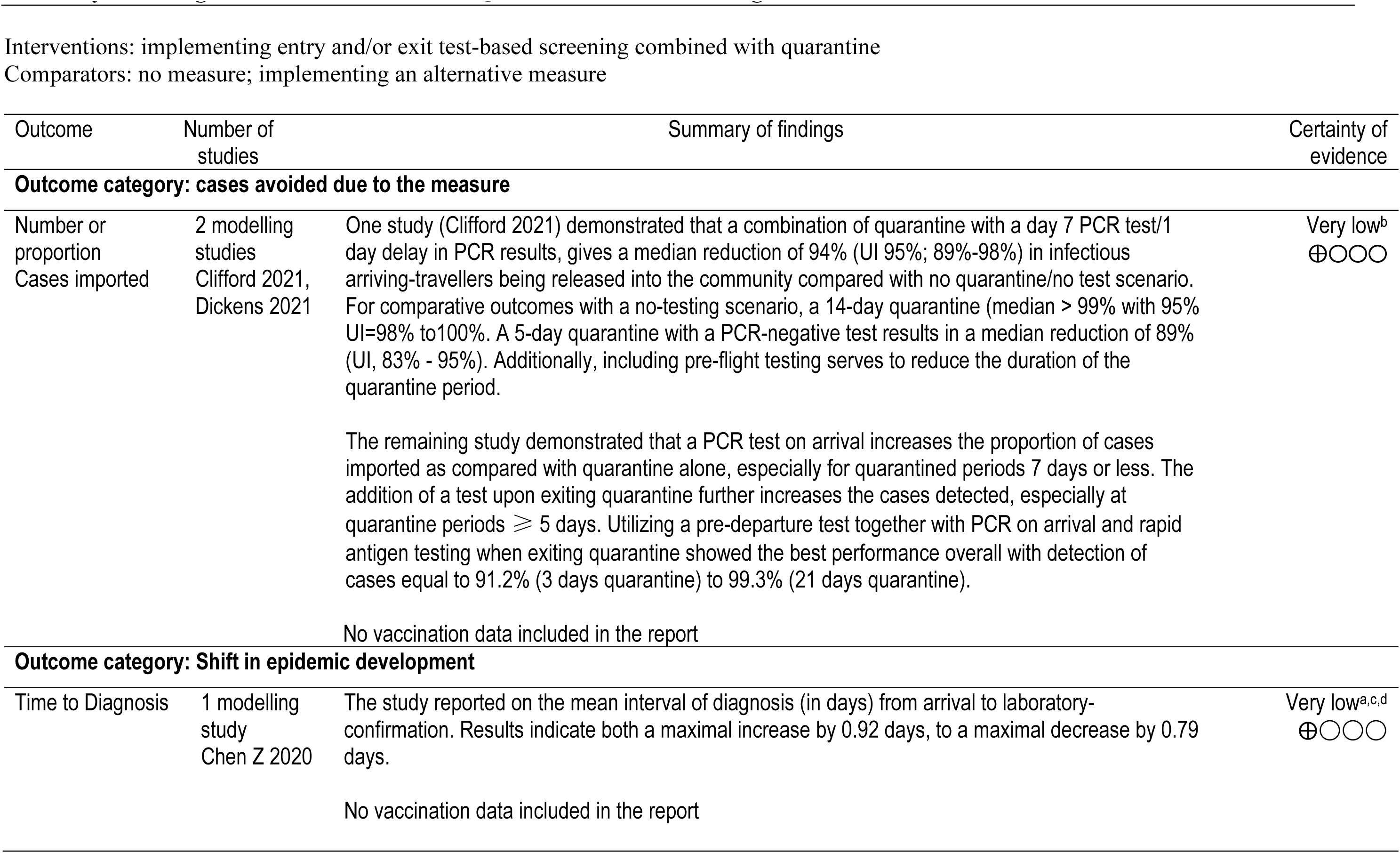
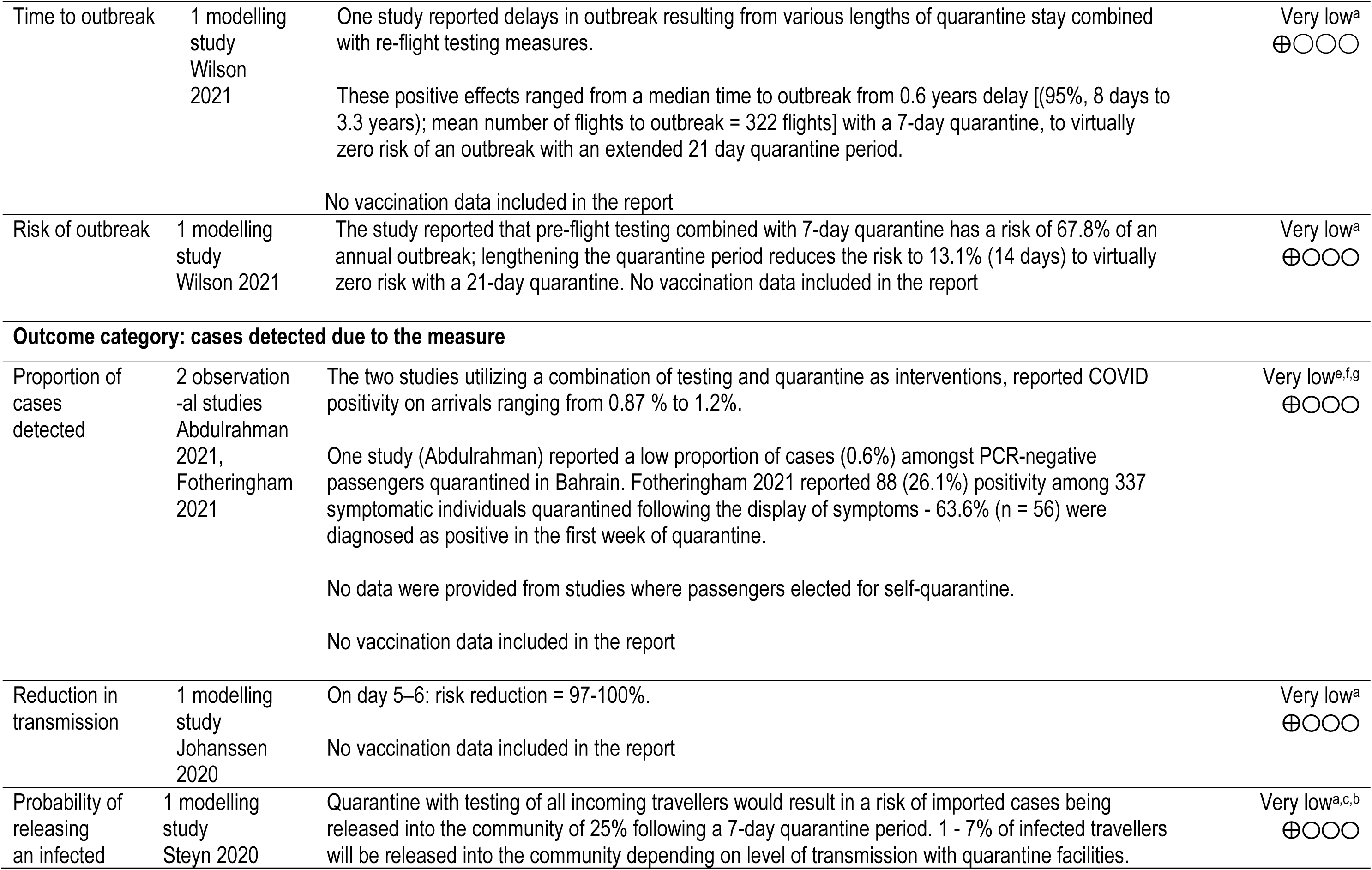
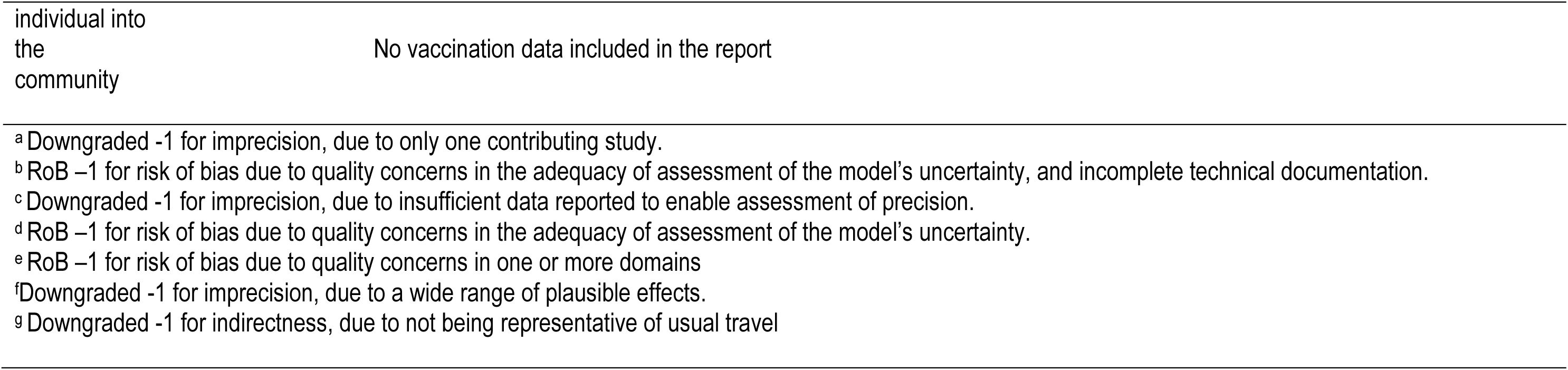
Combination of Quarantine and Microbiological Detection

**Table 4.**
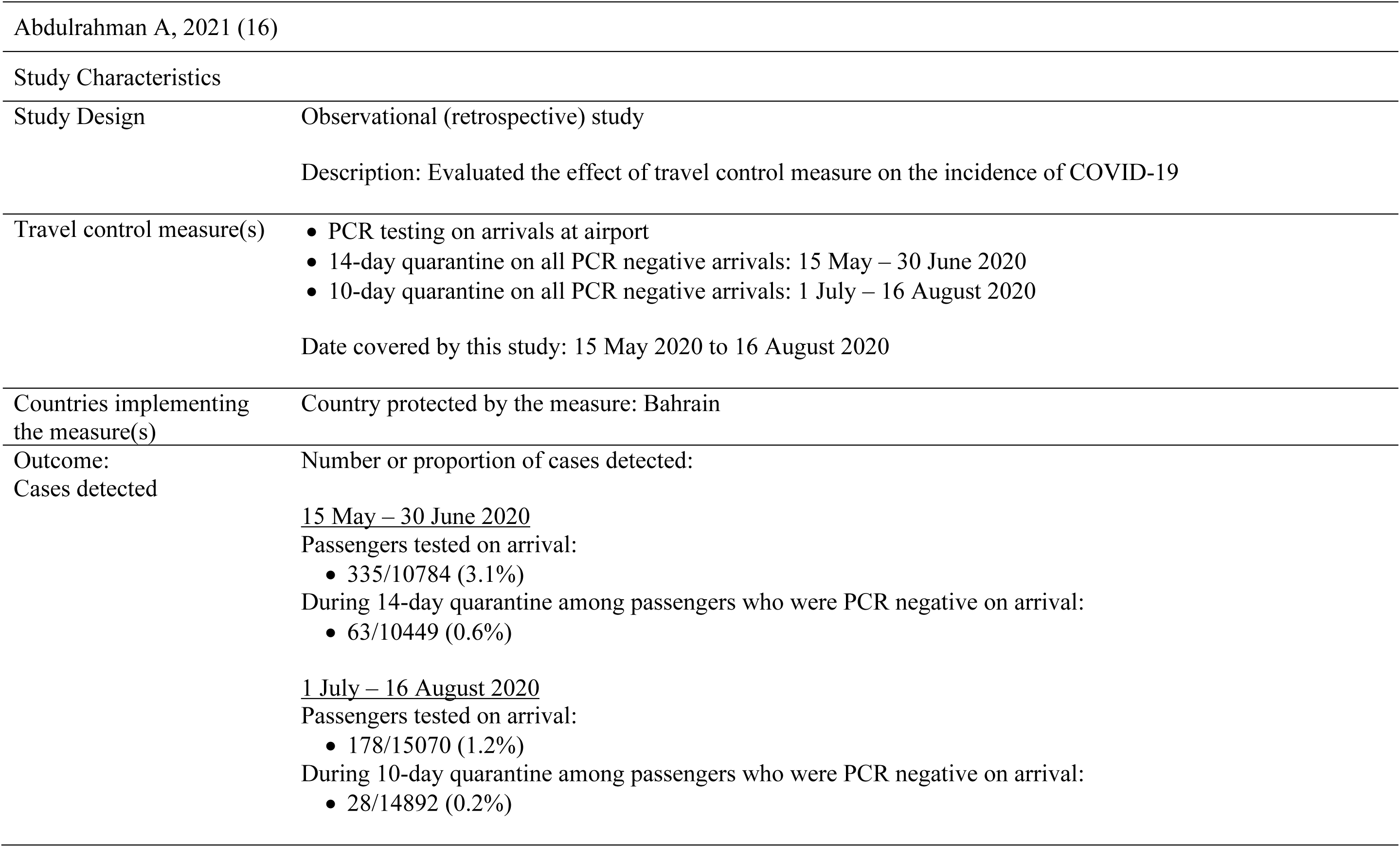
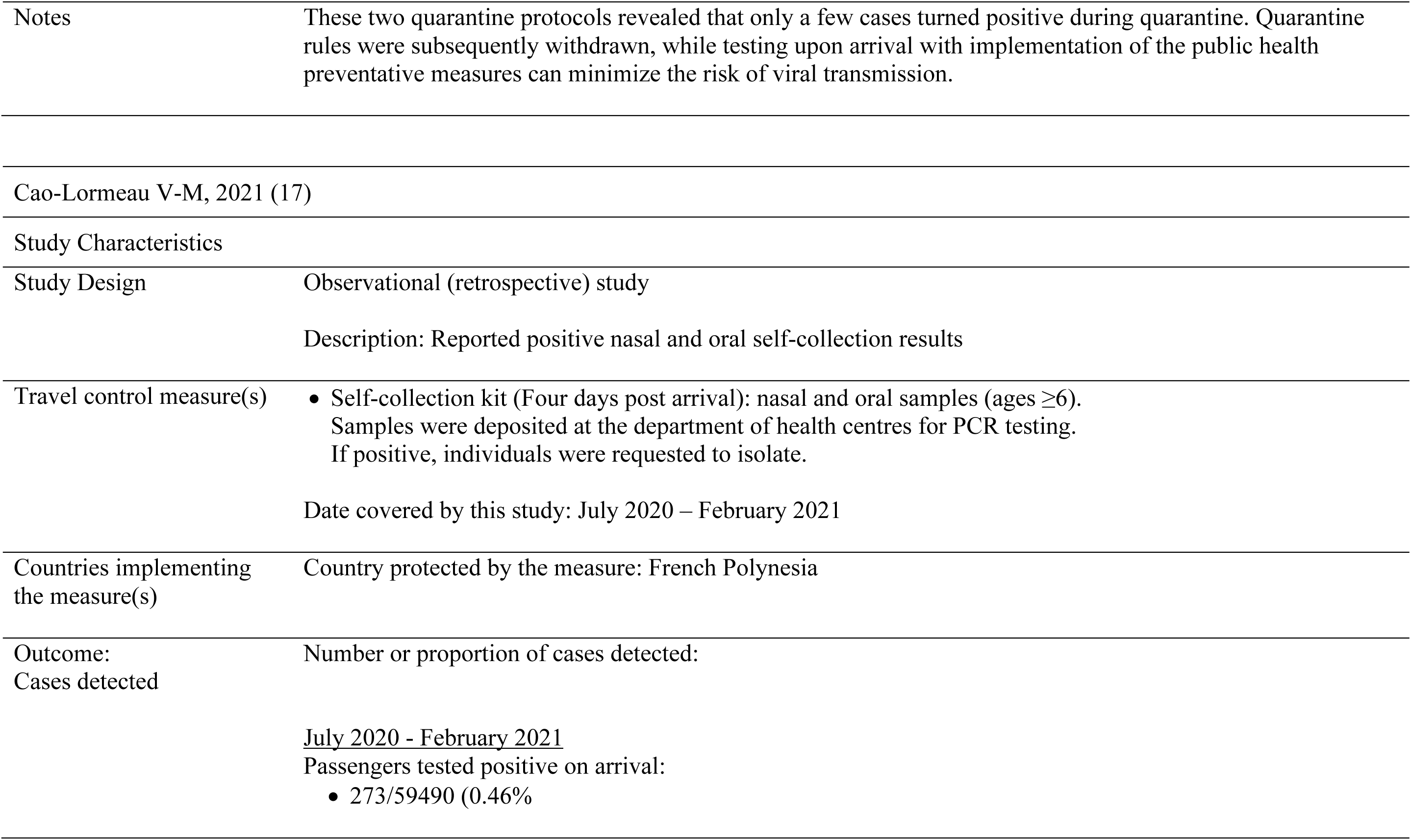
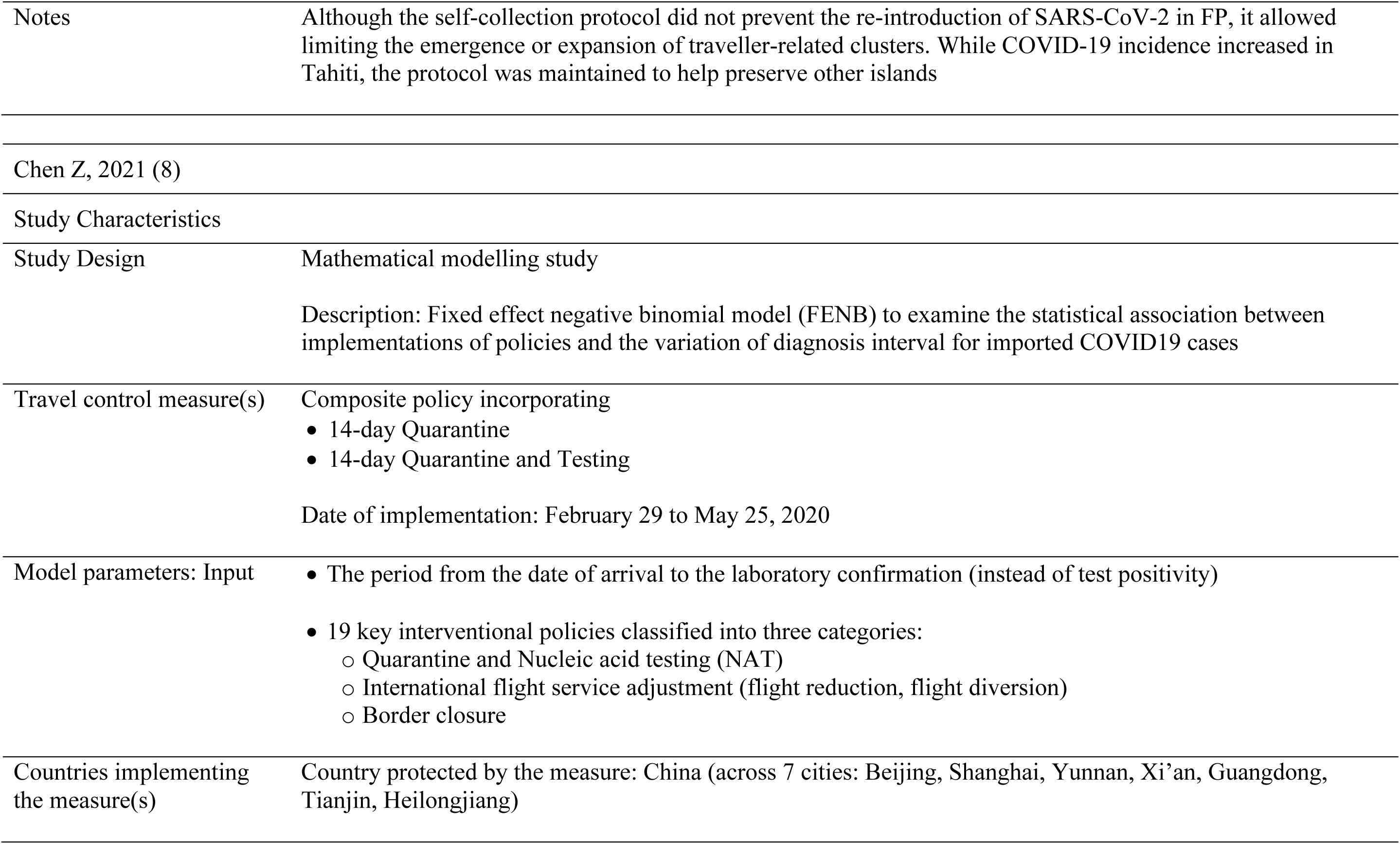
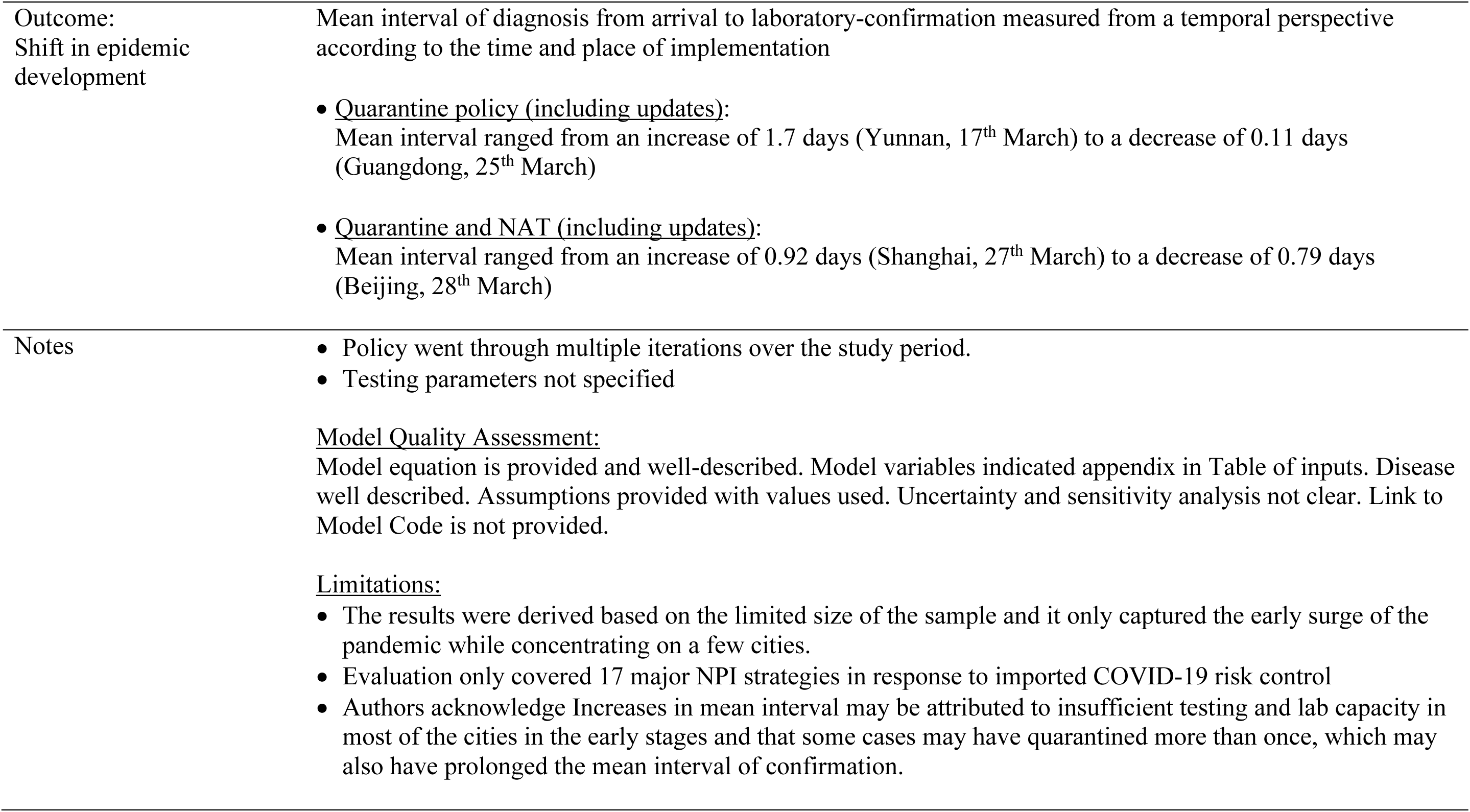
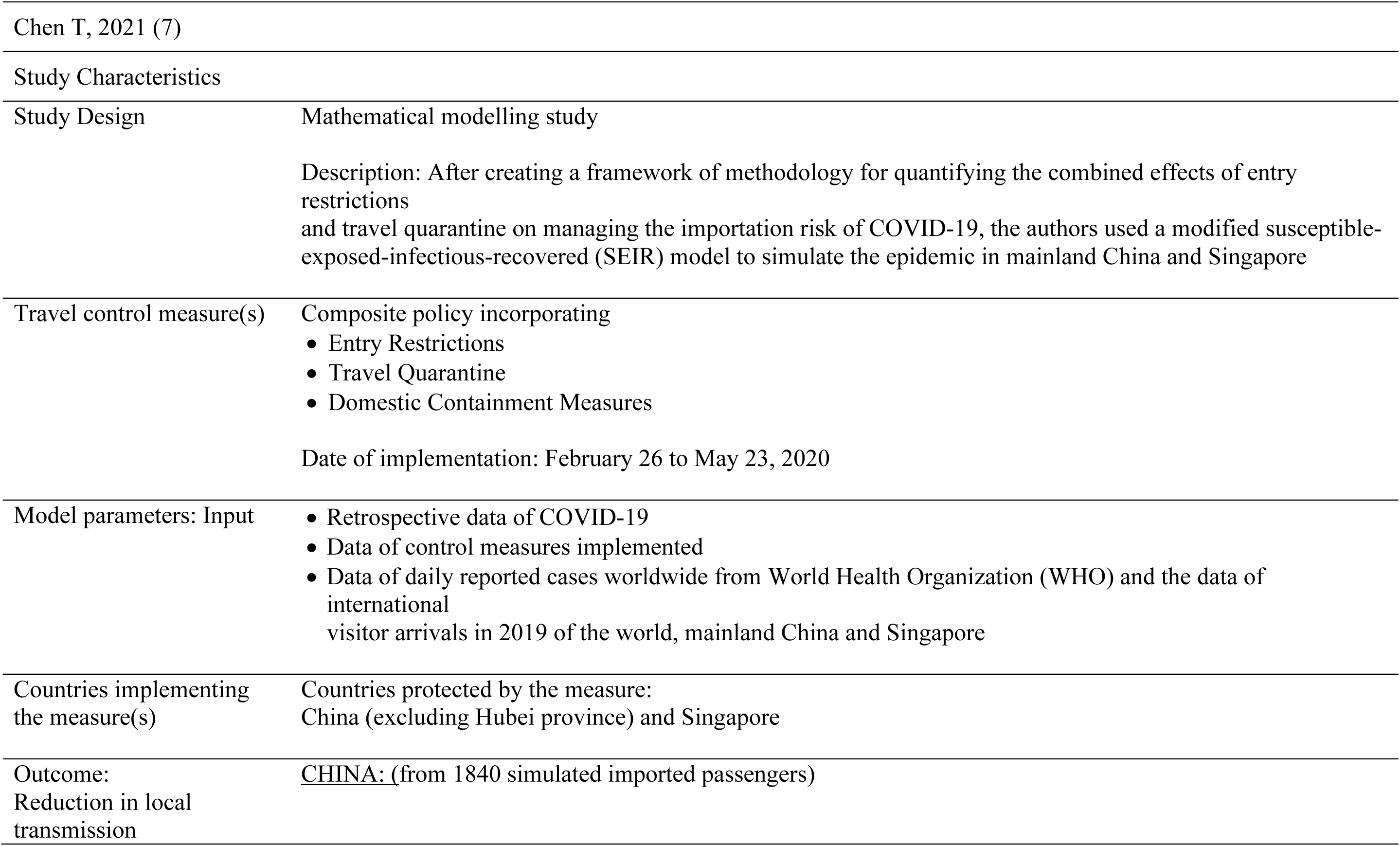
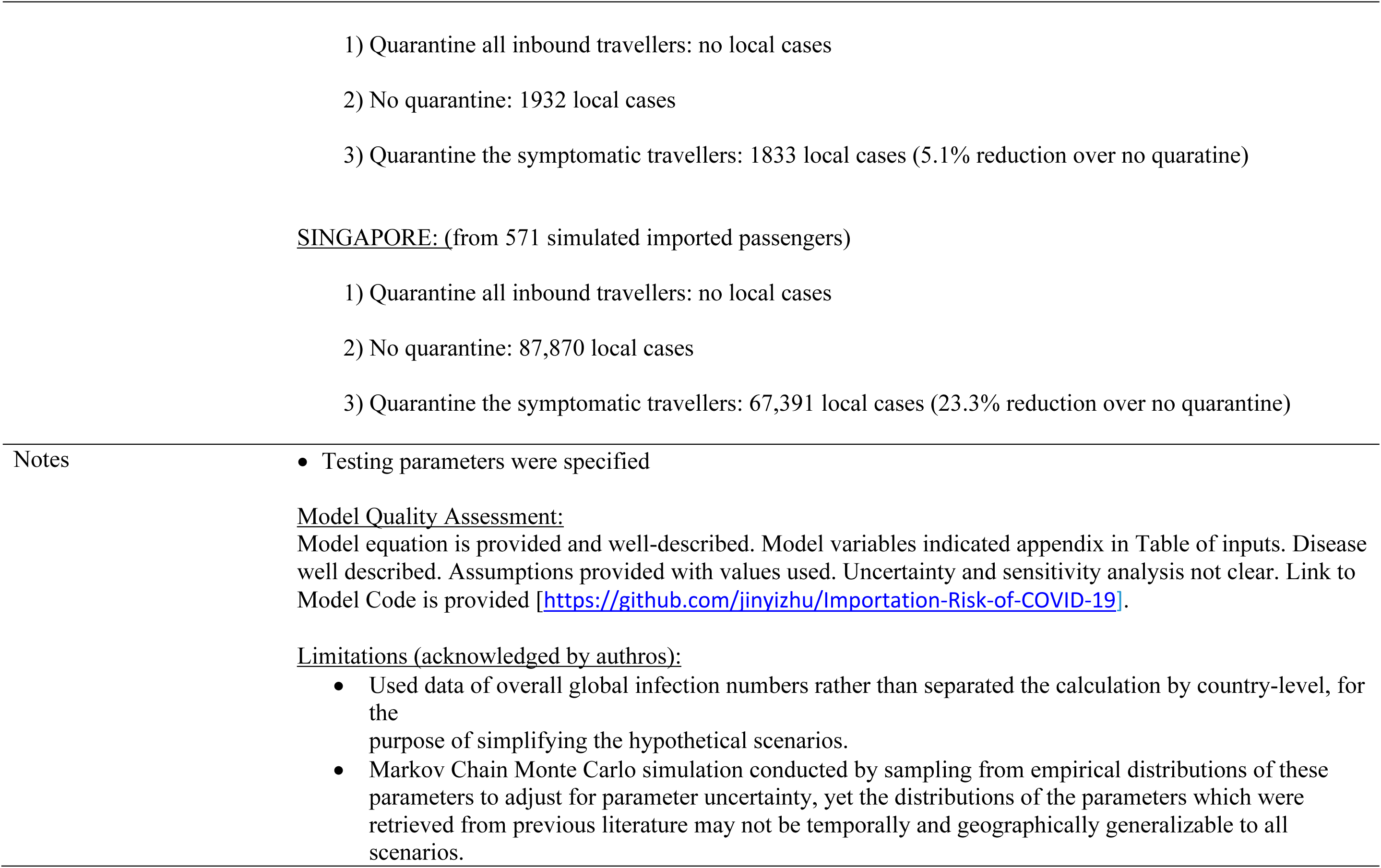
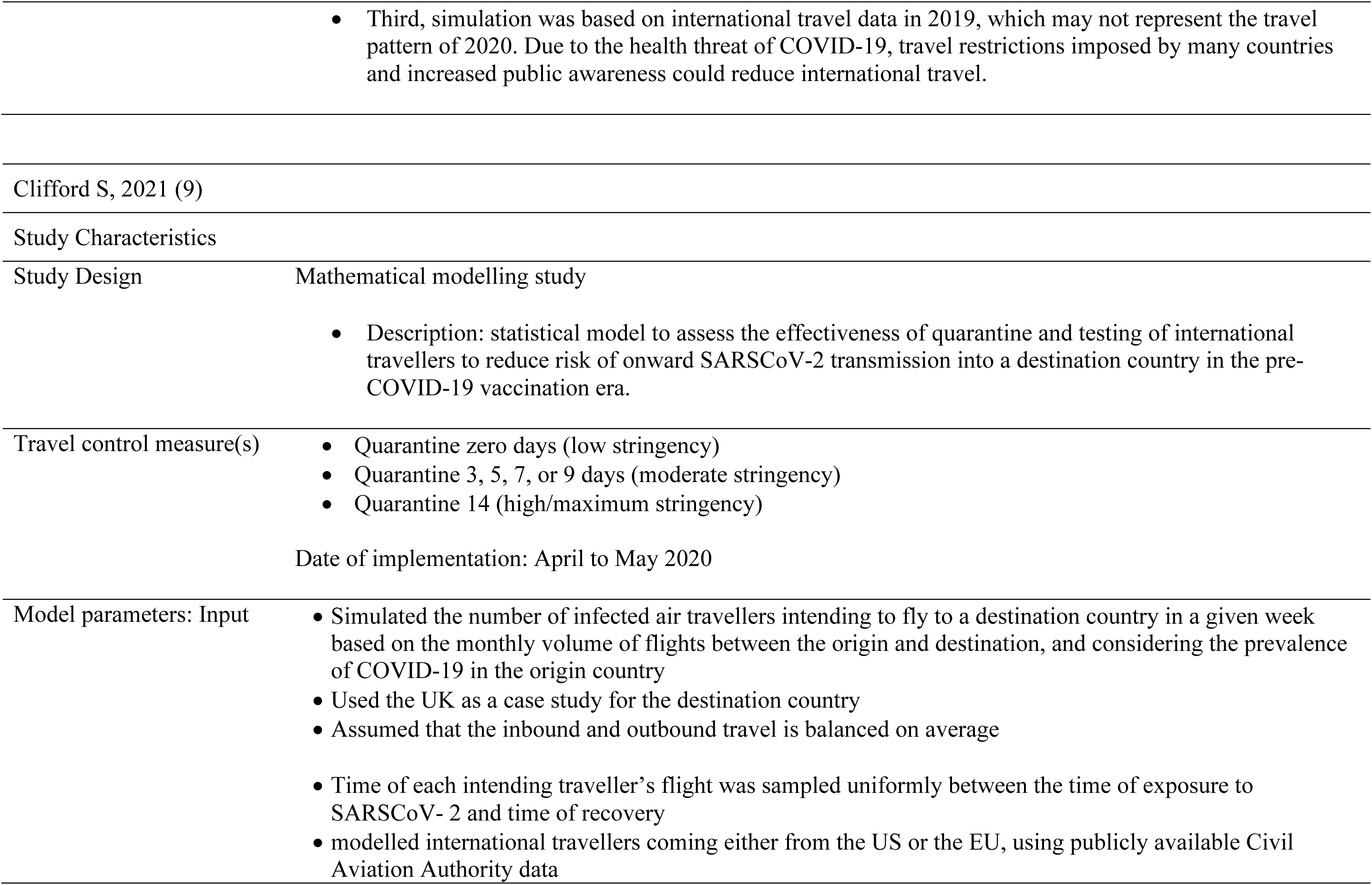
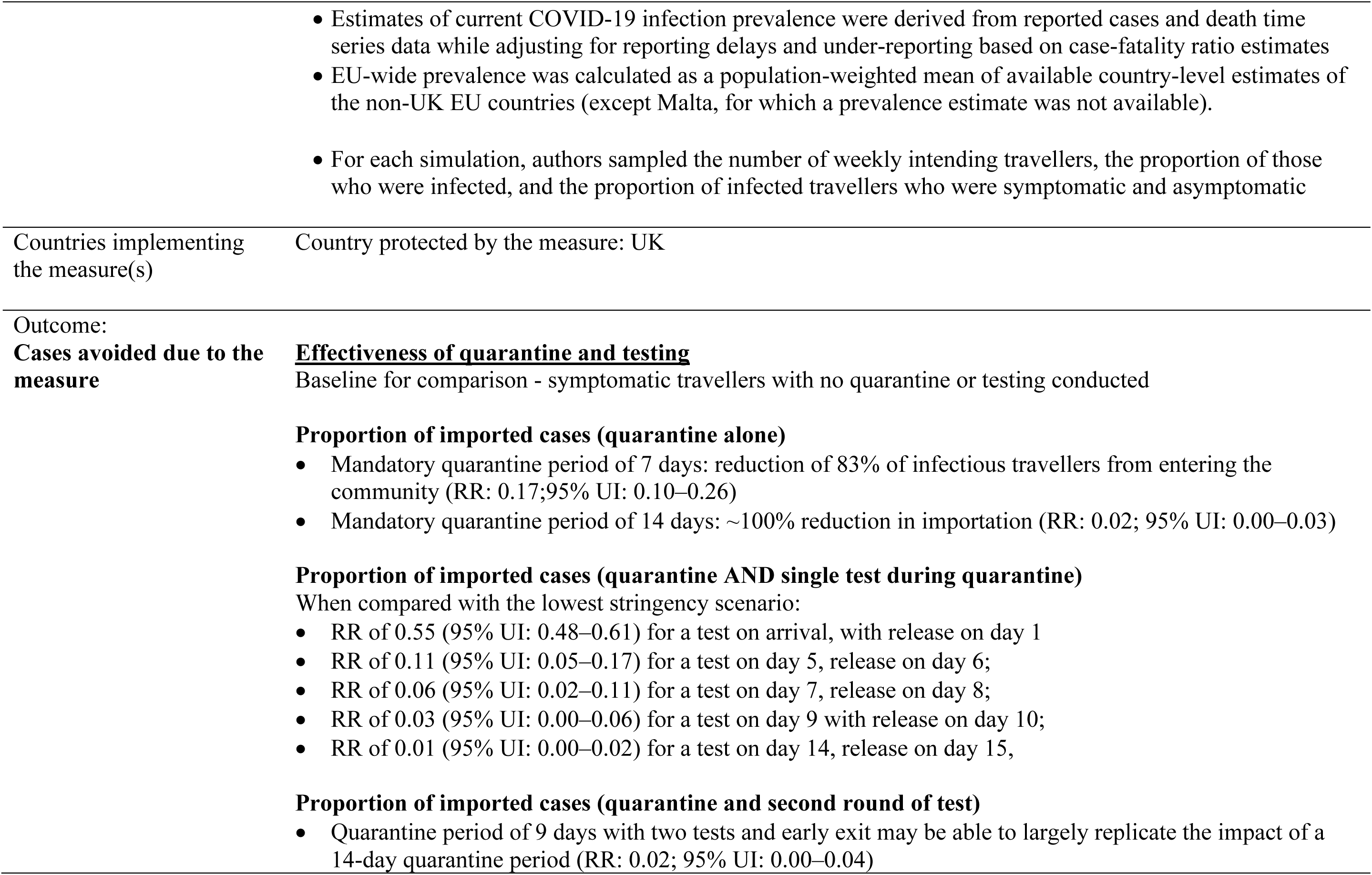
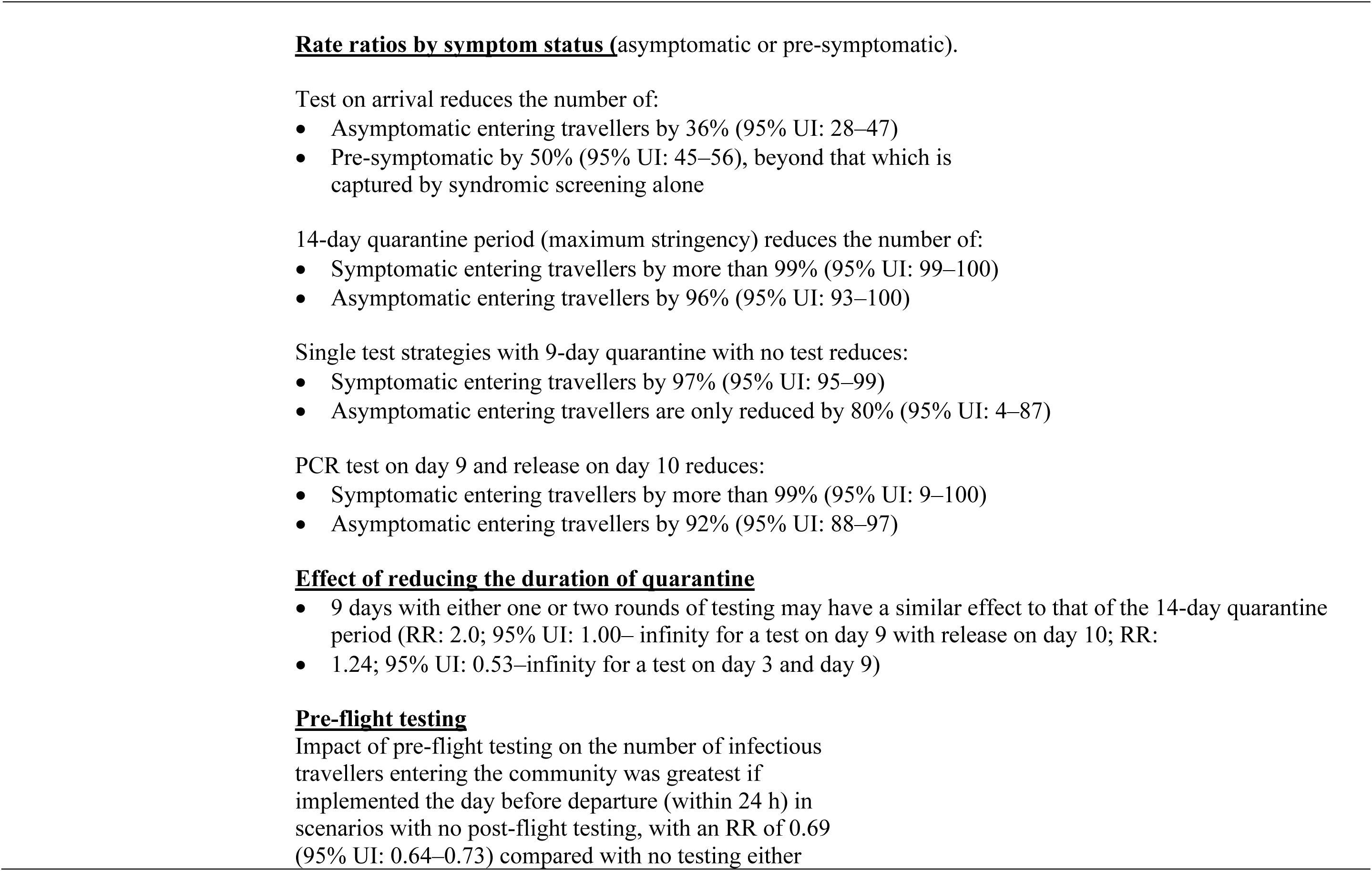
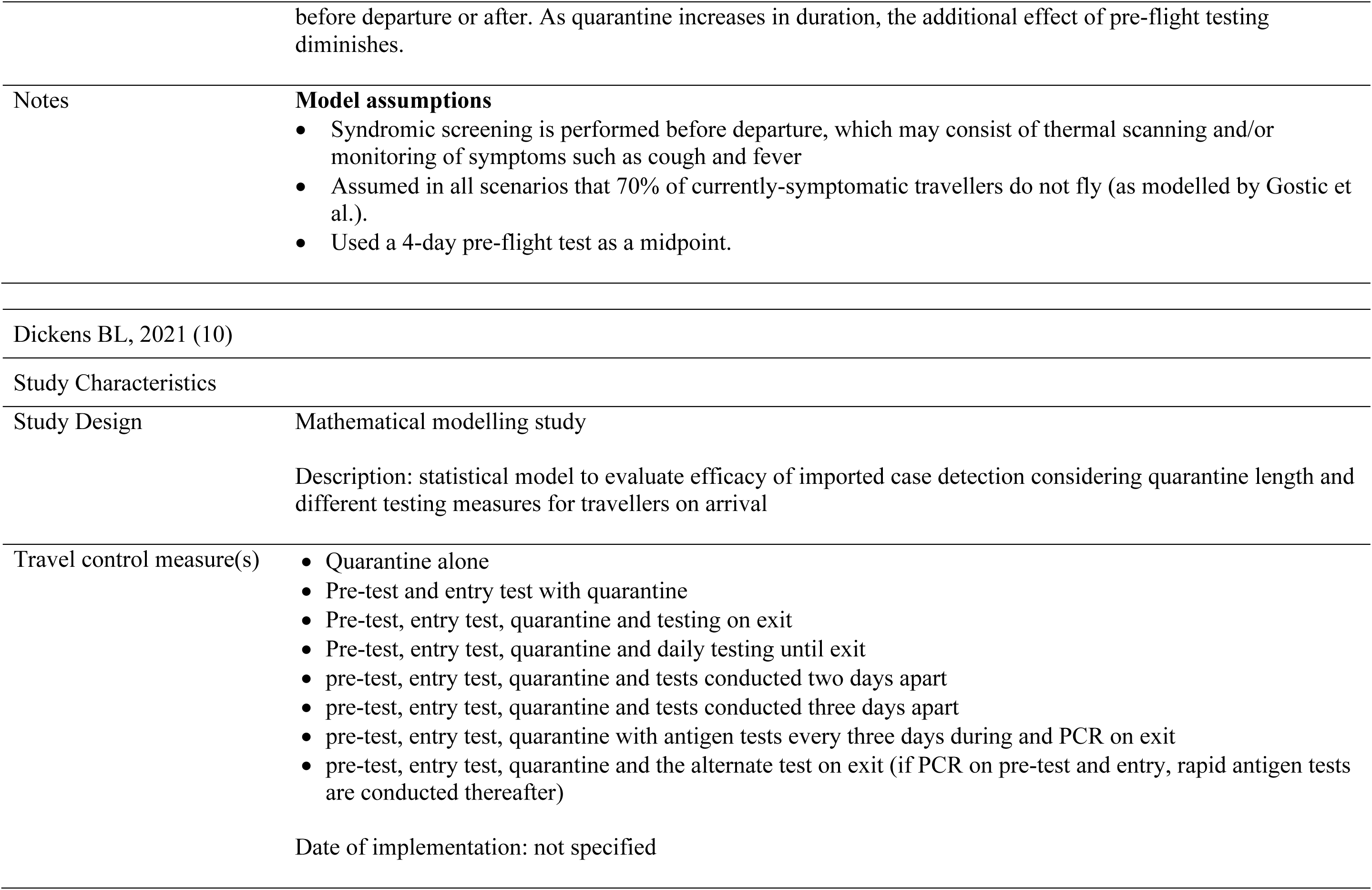
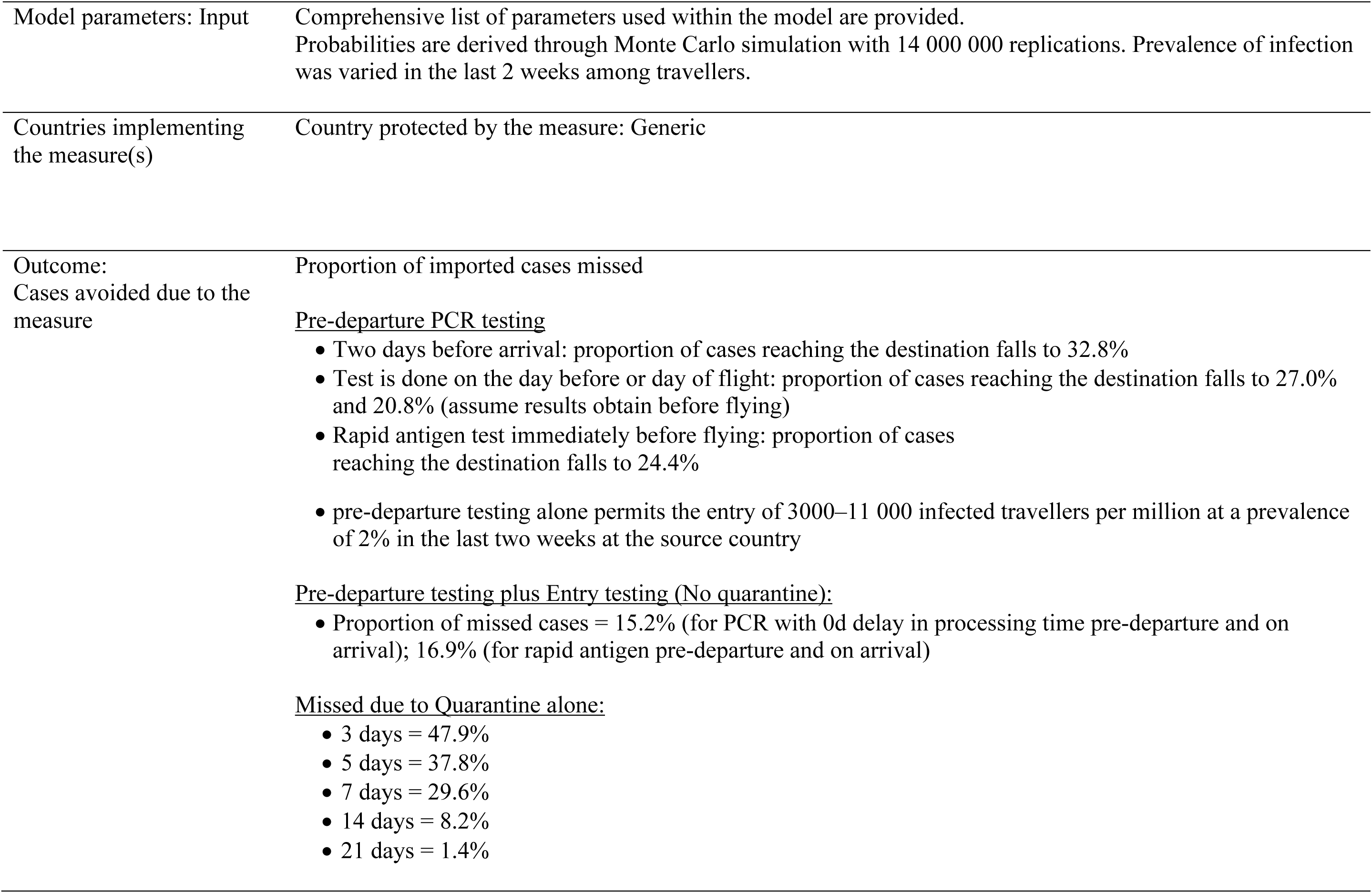
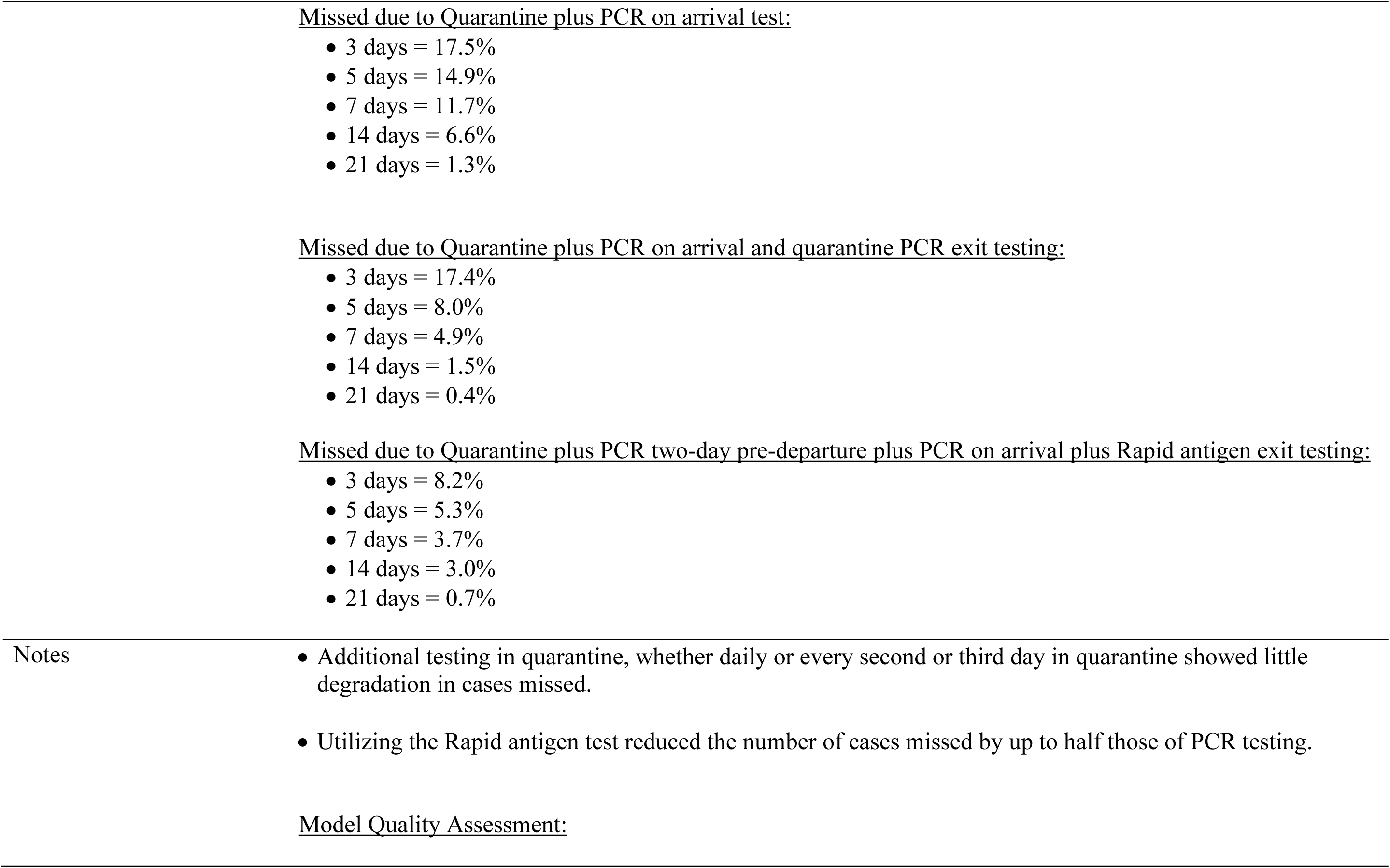
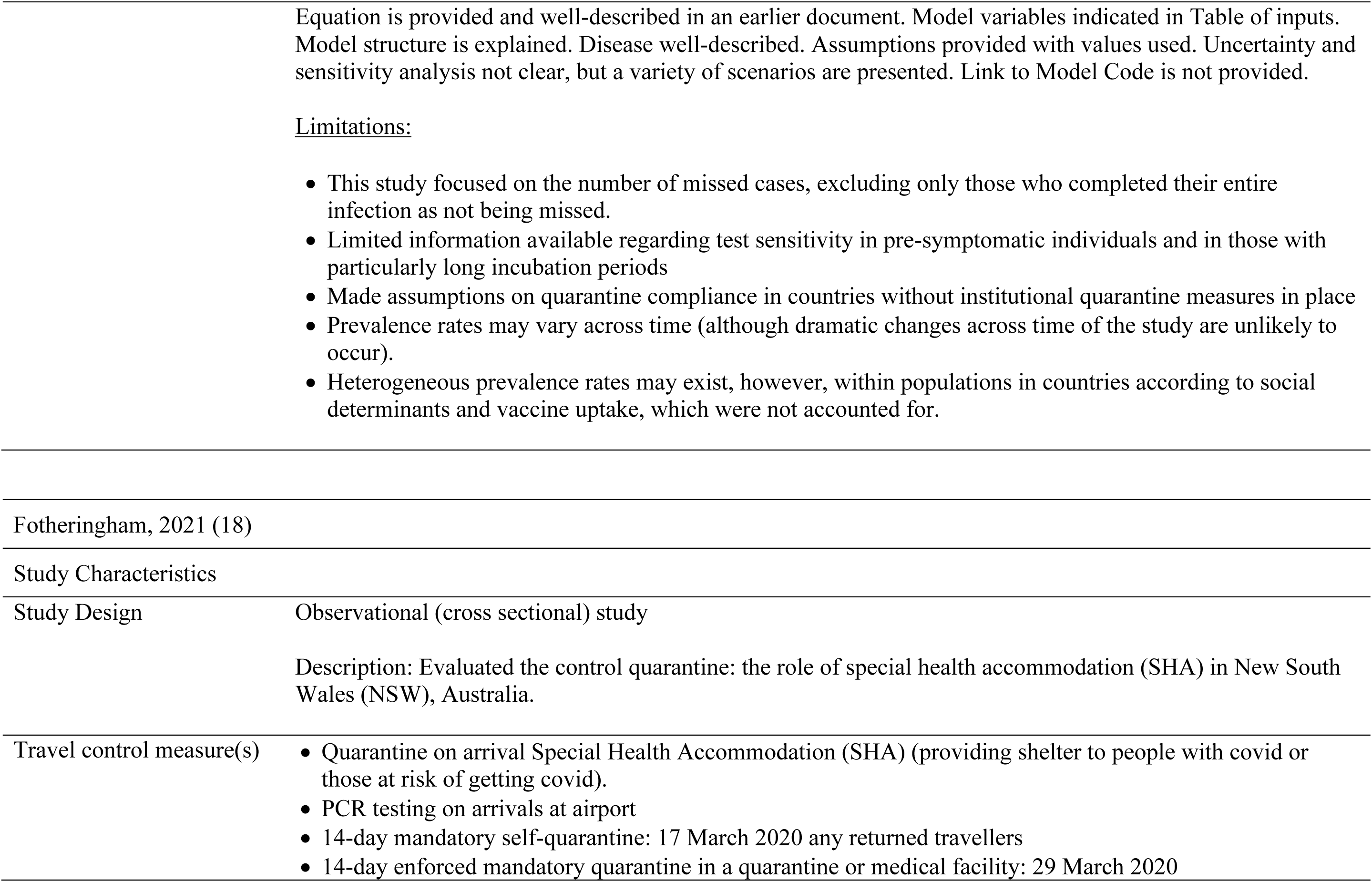
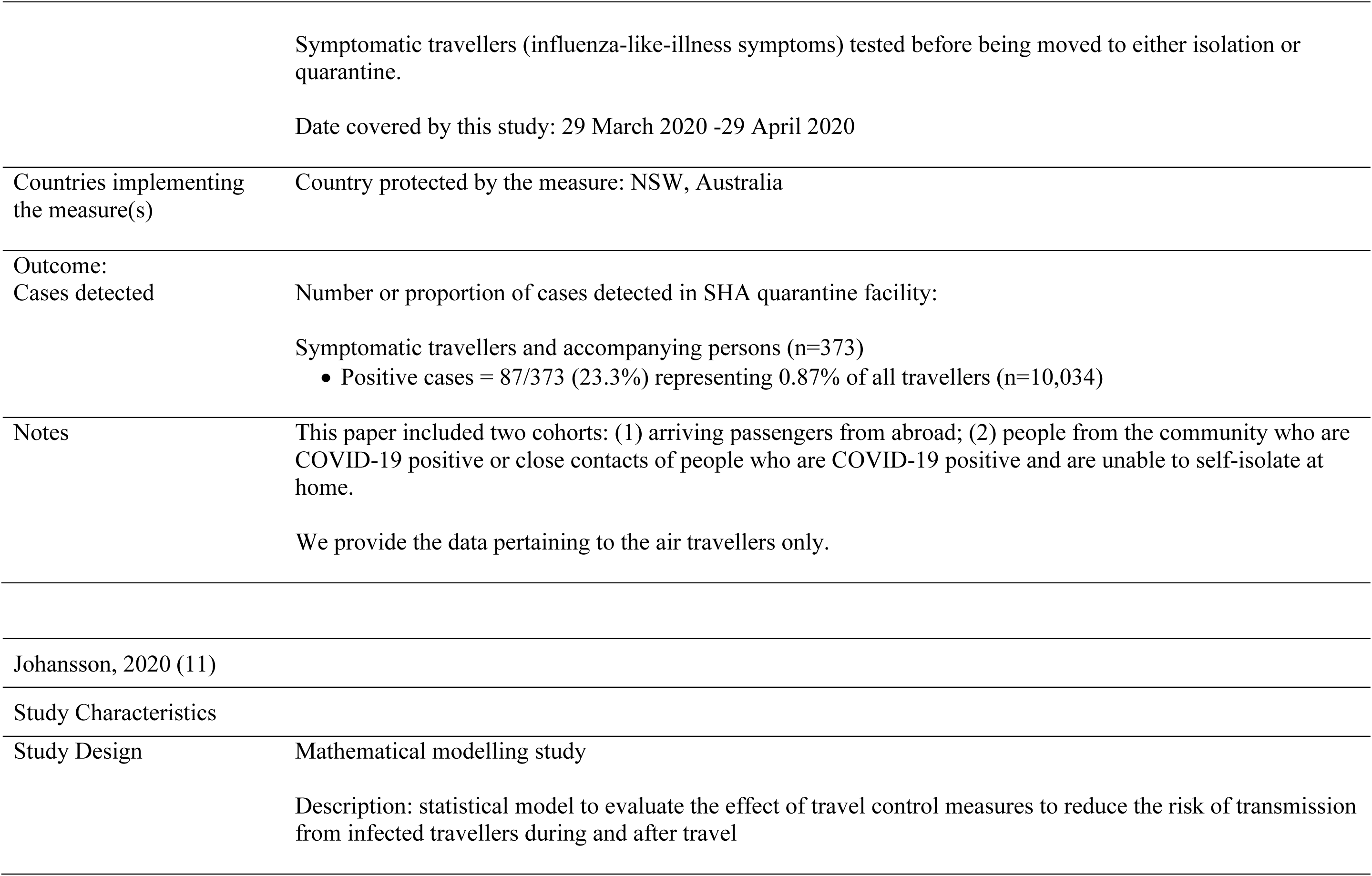
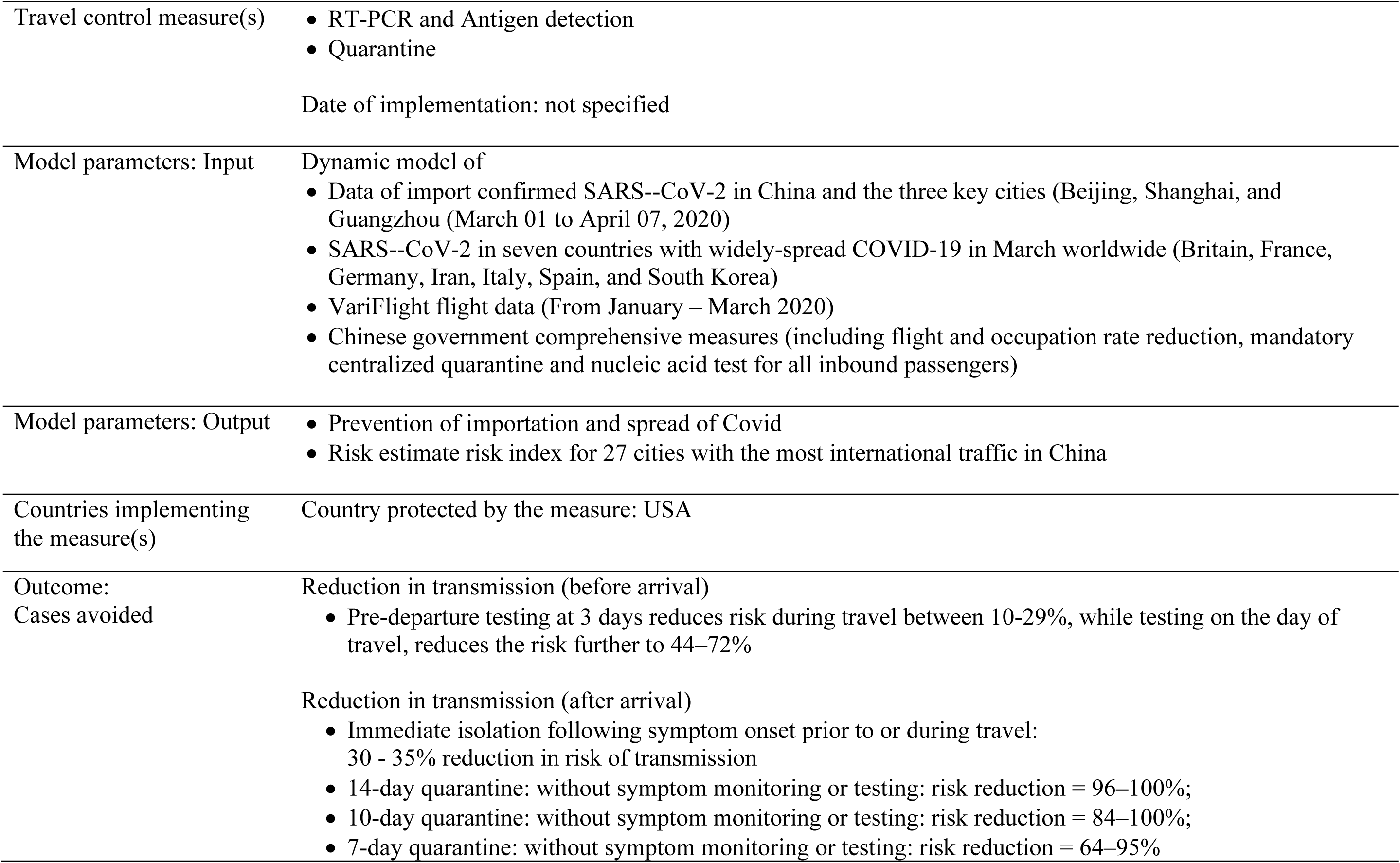
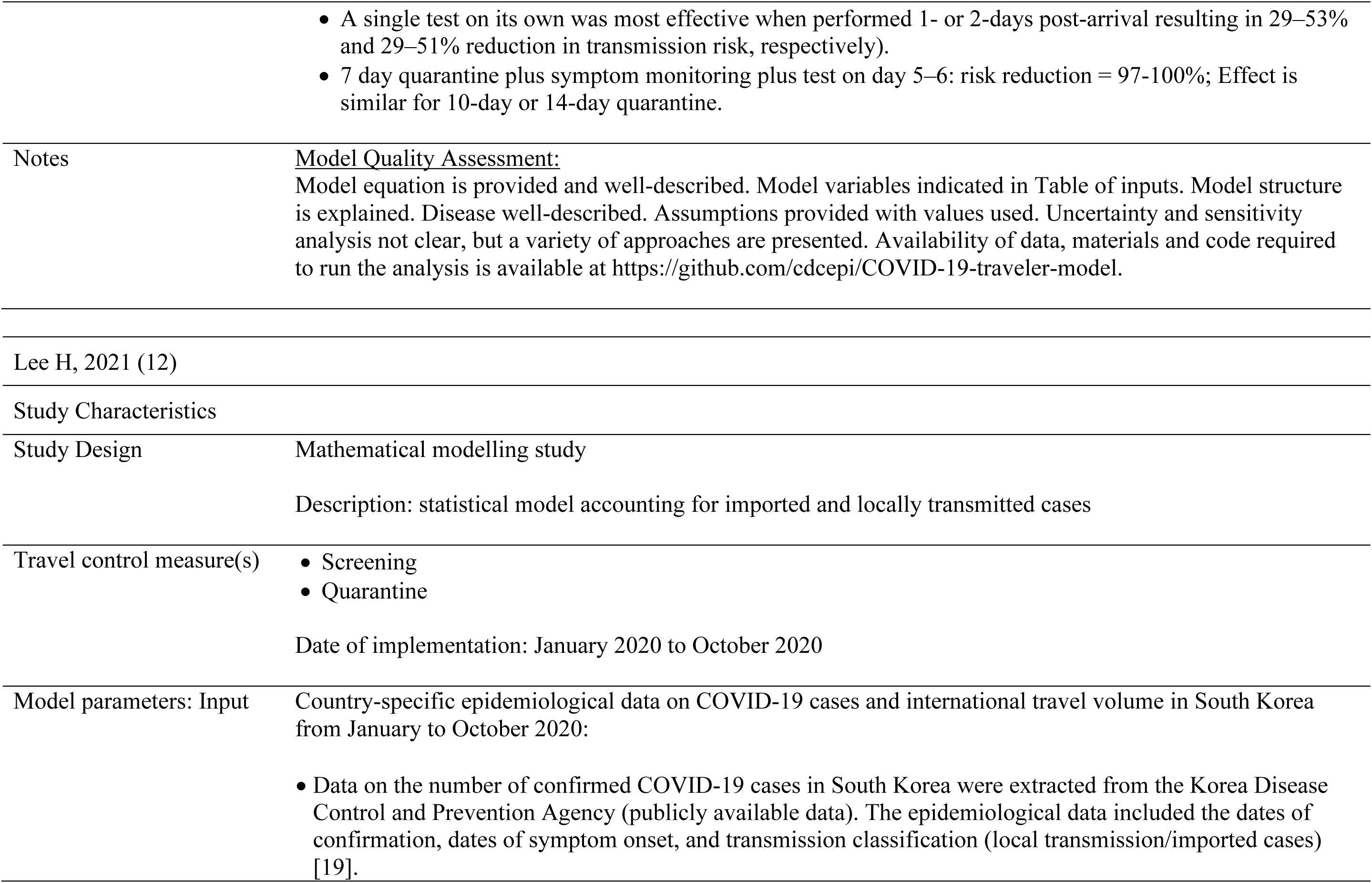
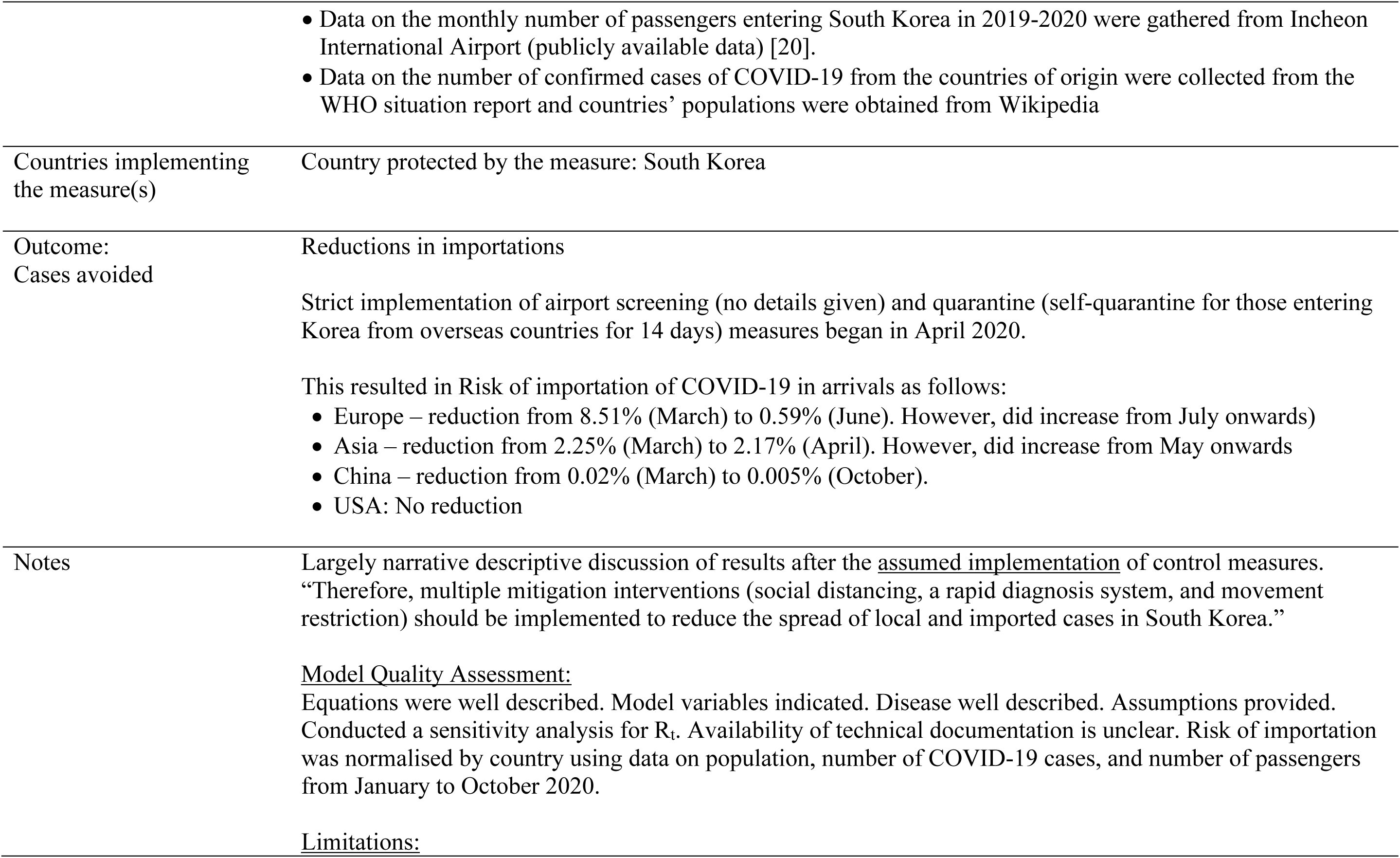
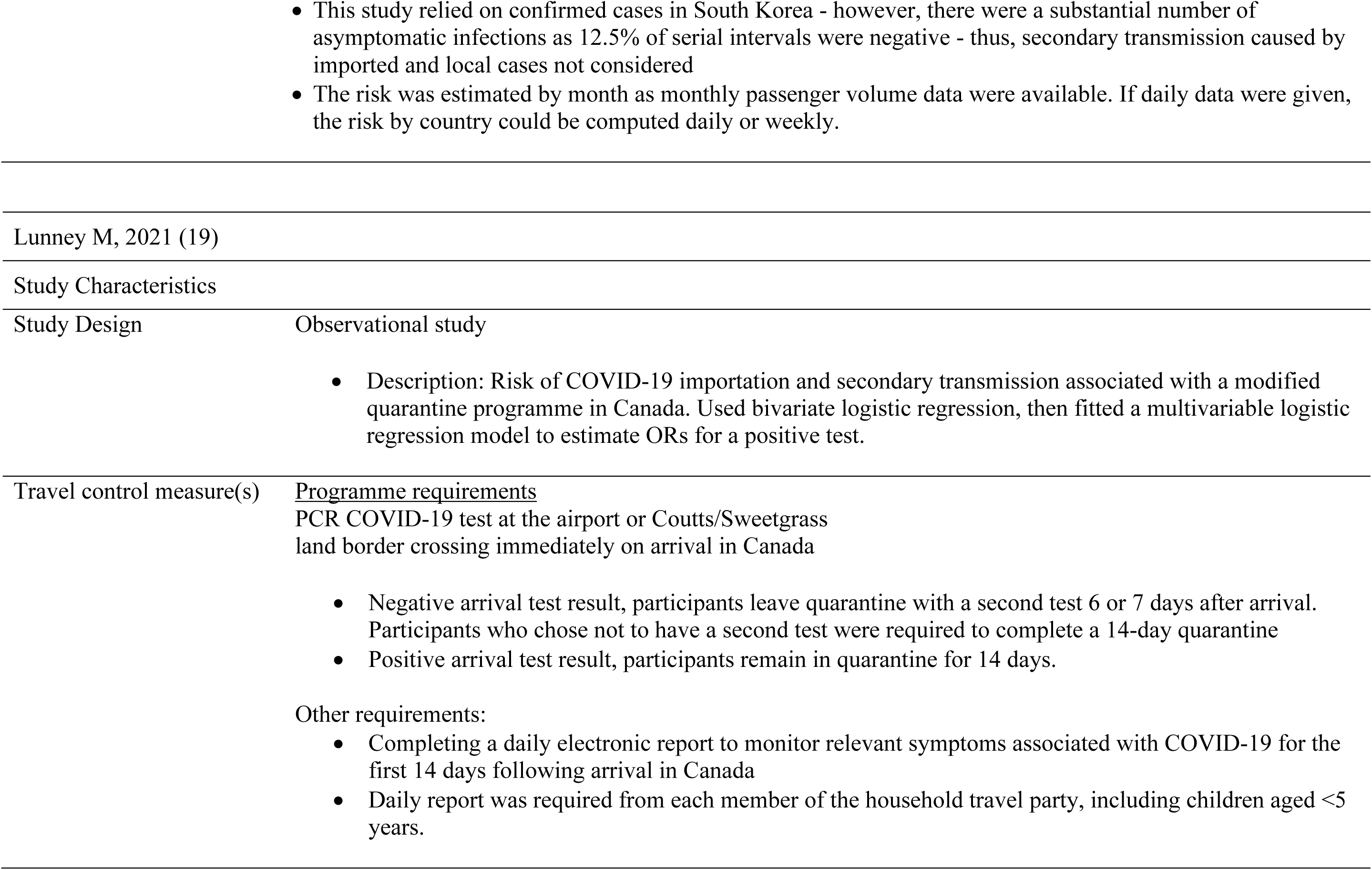
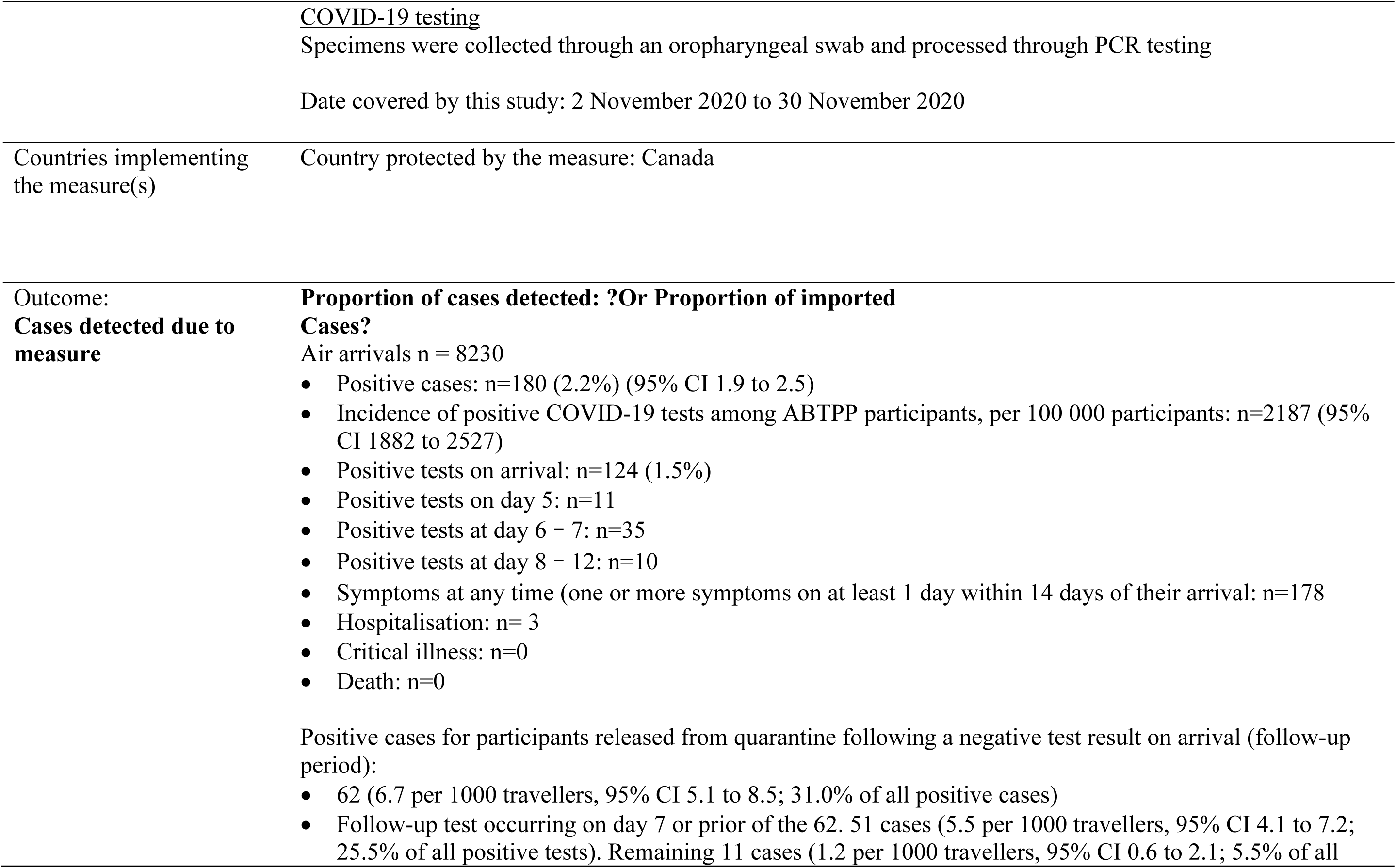
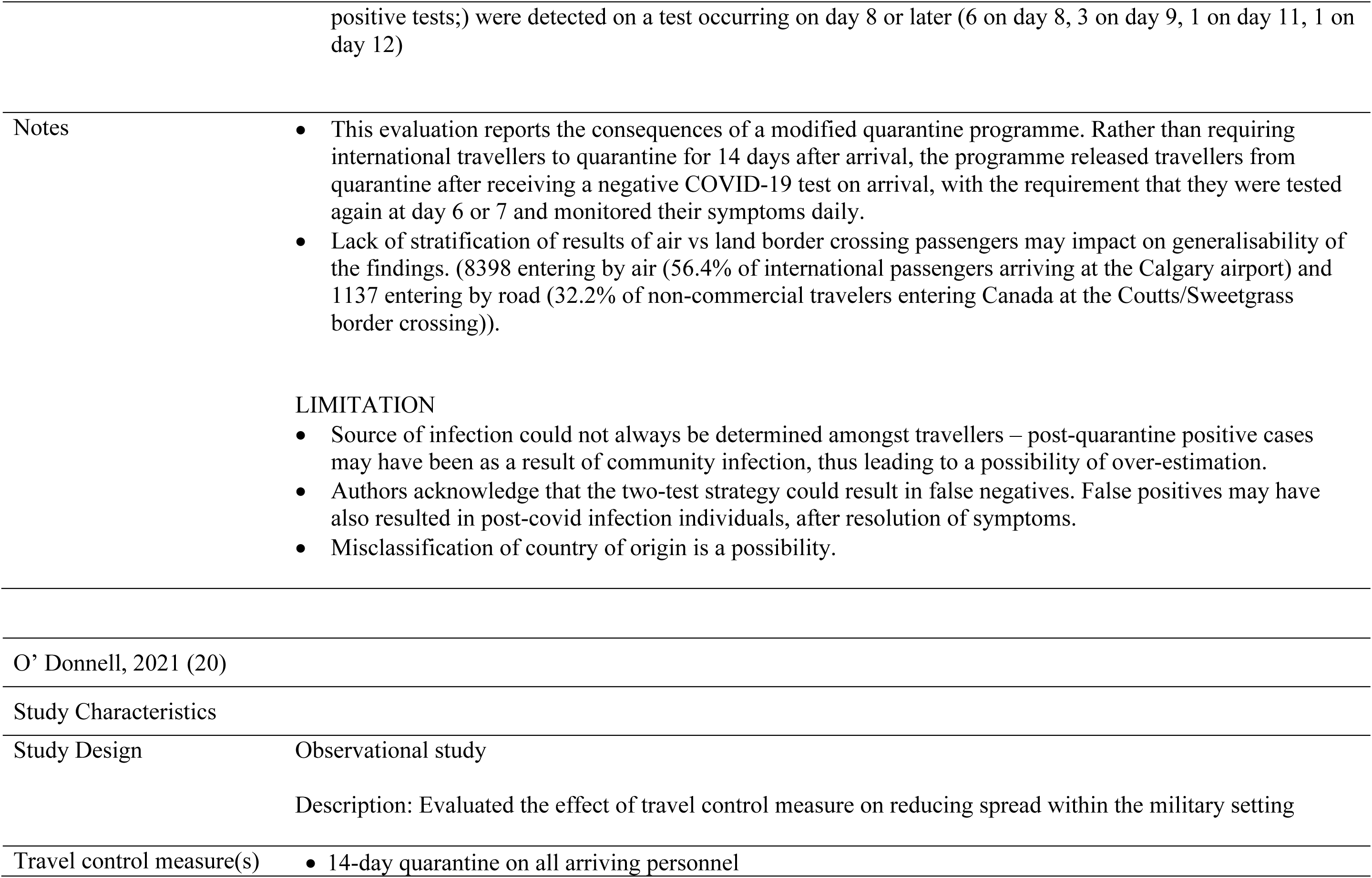
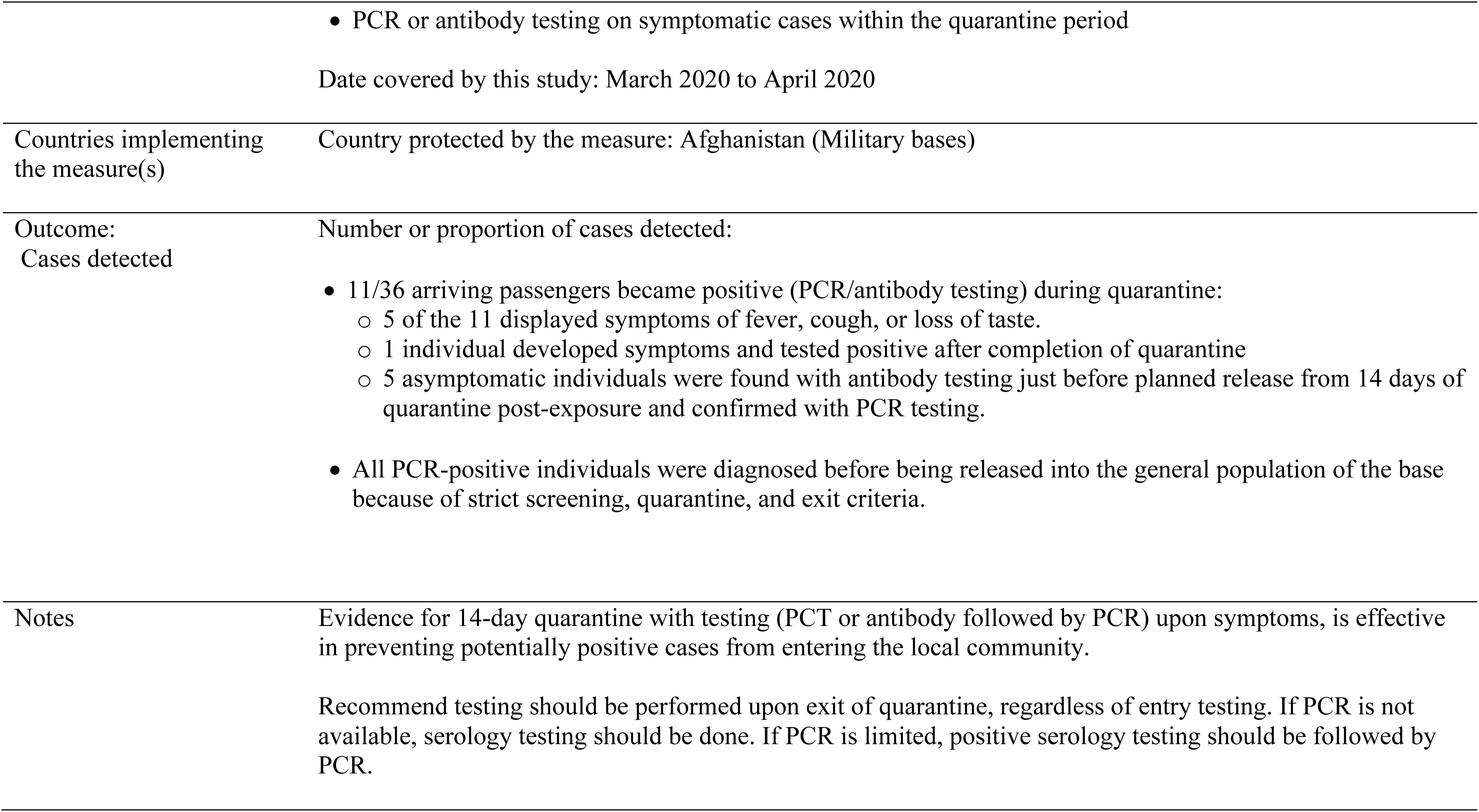
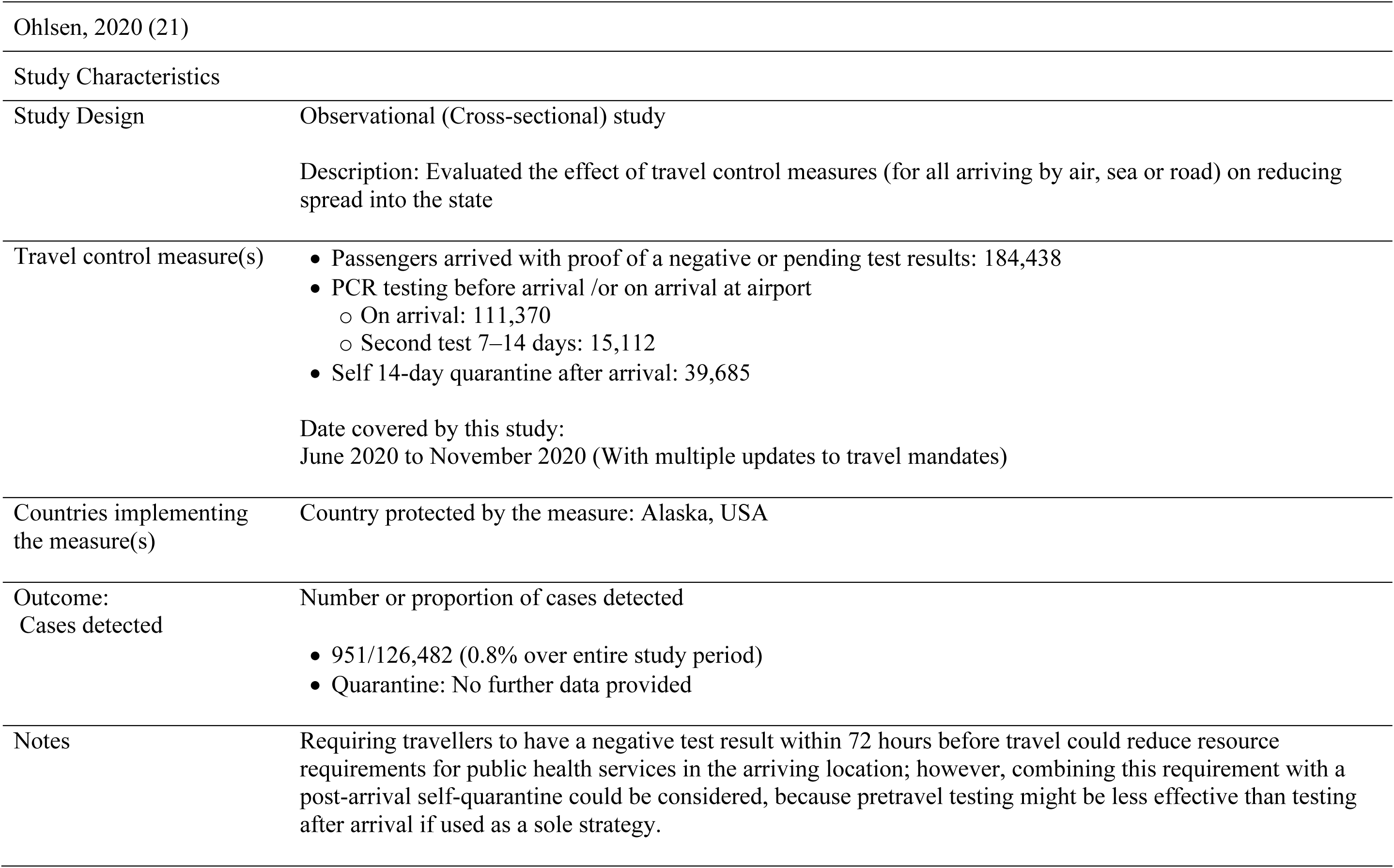
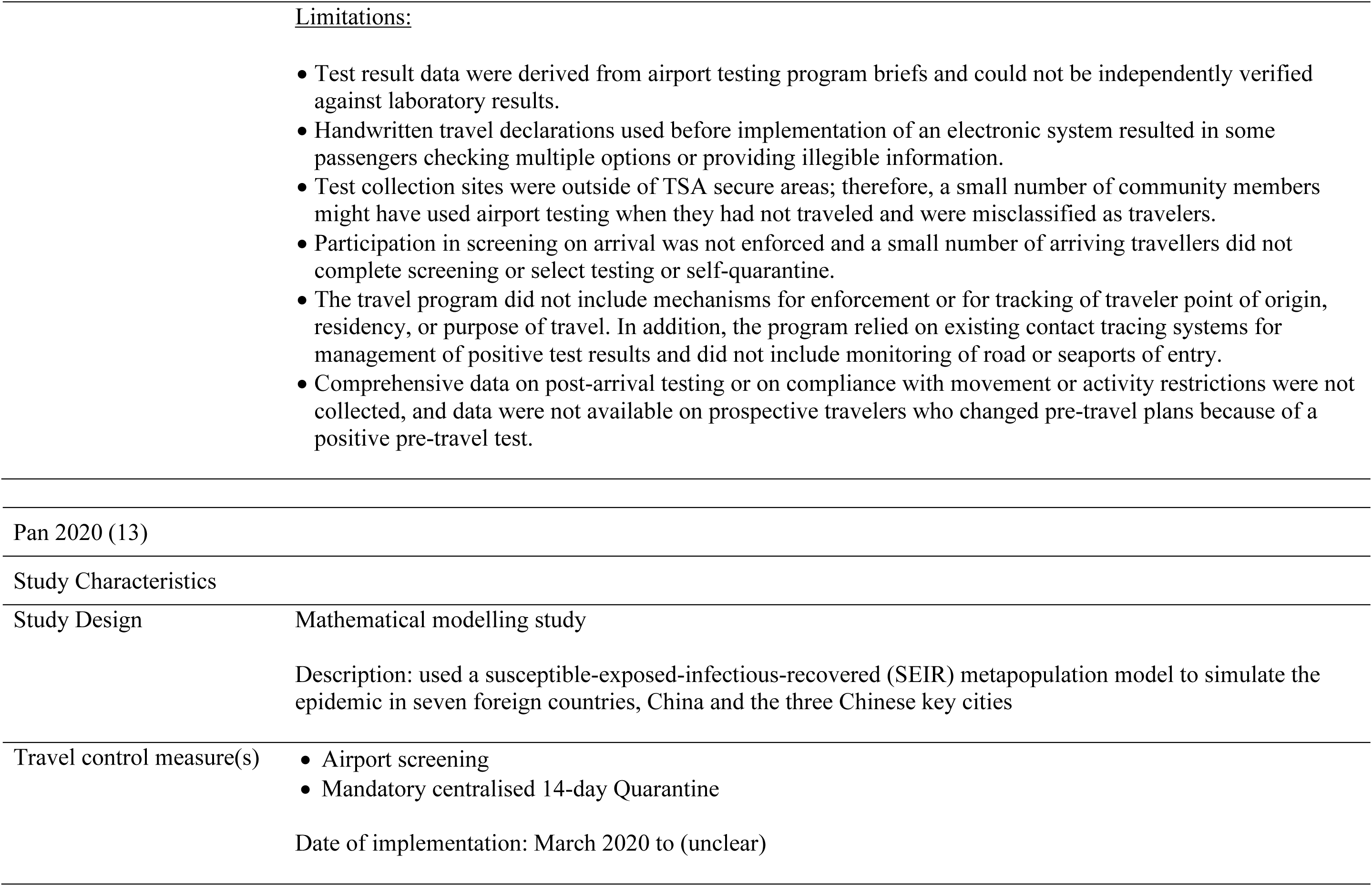
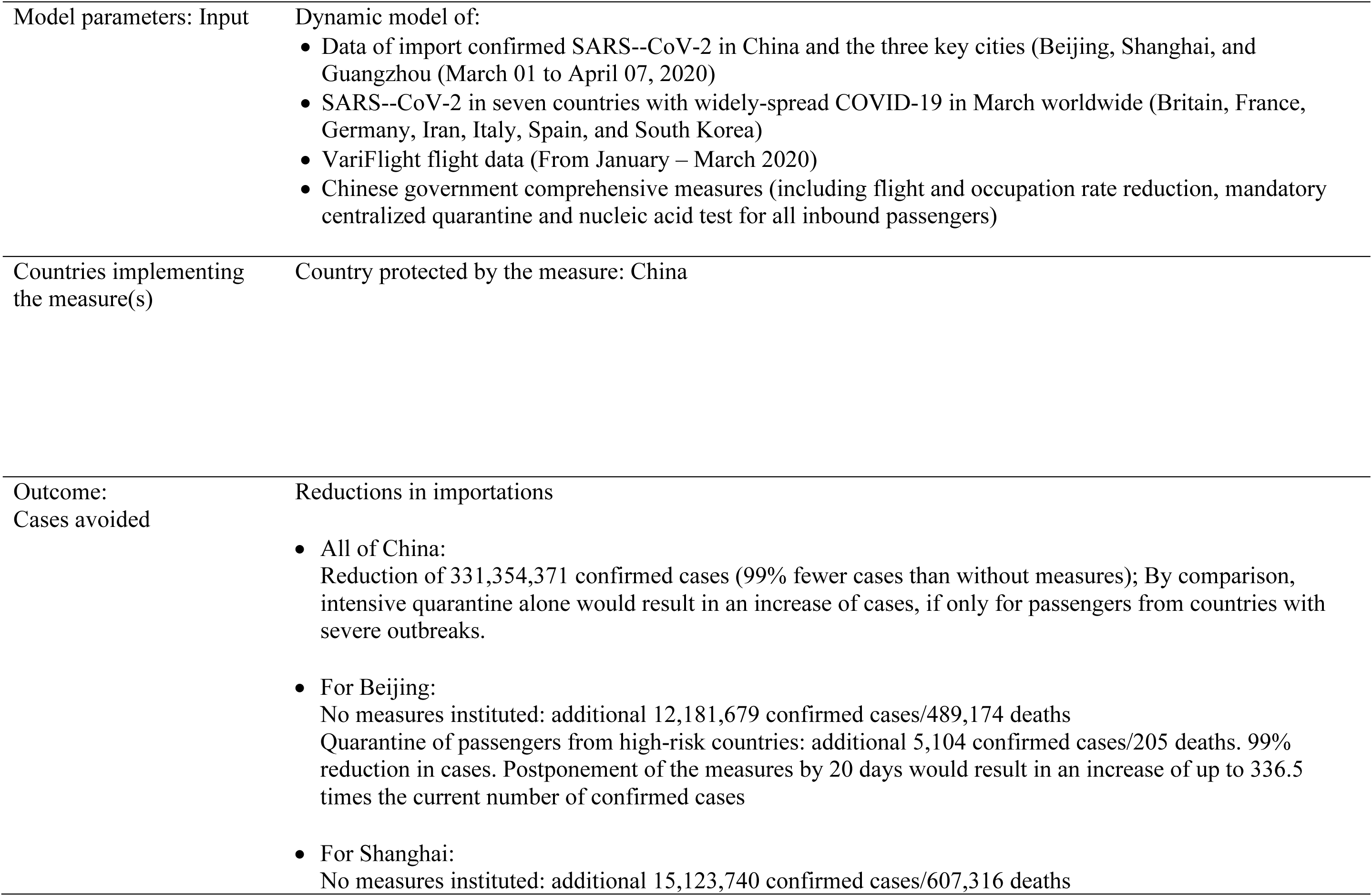
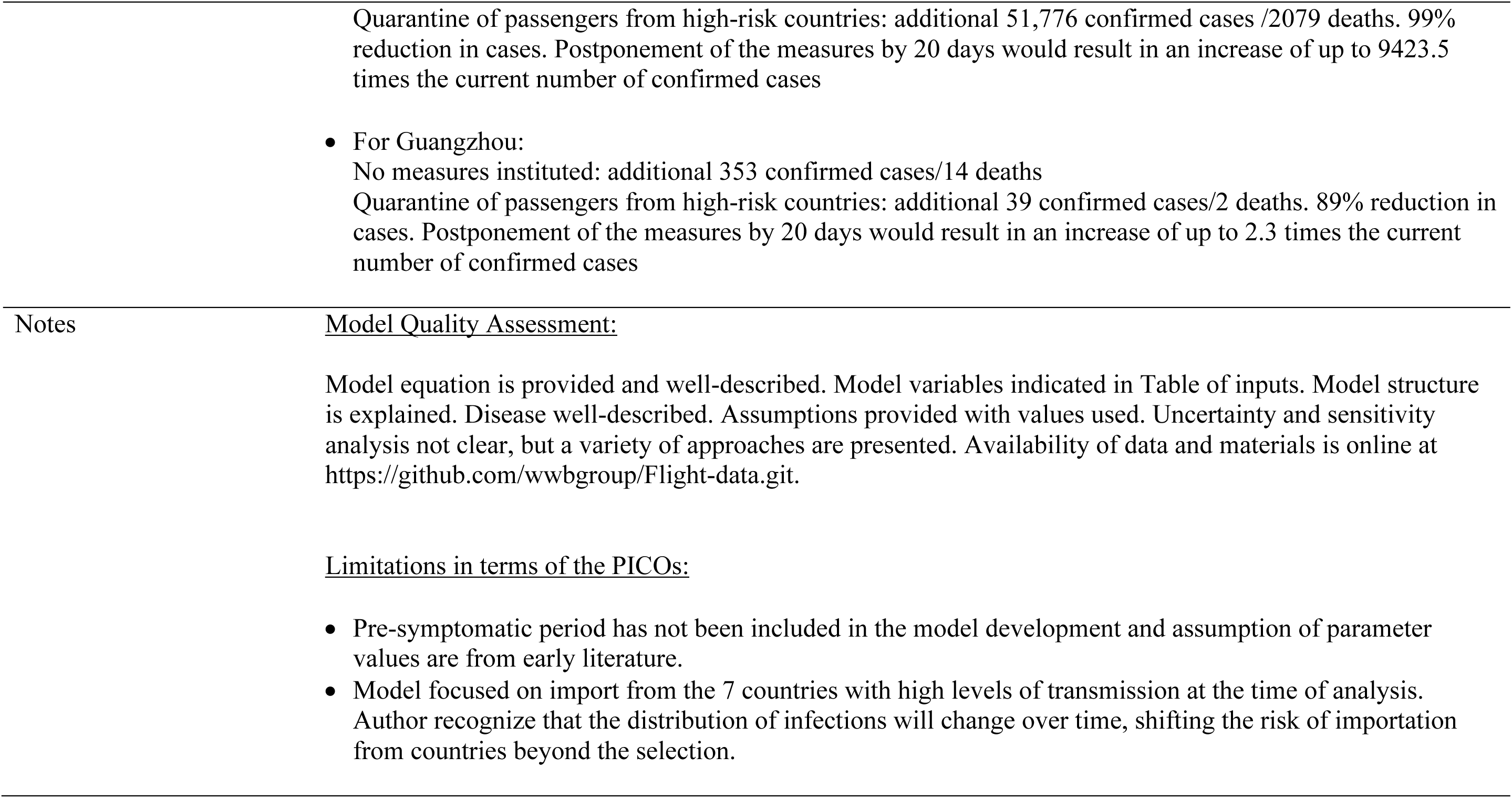
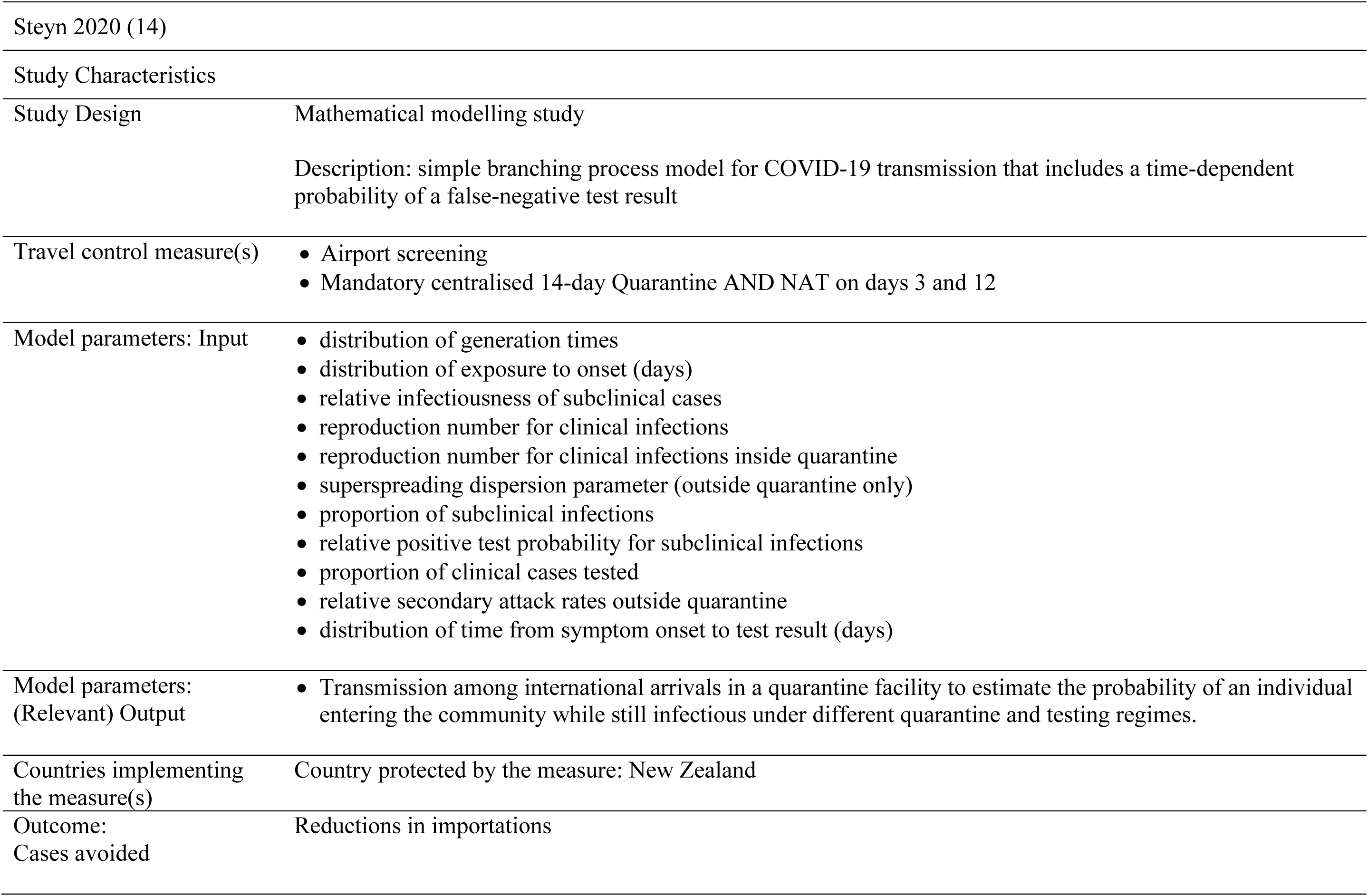
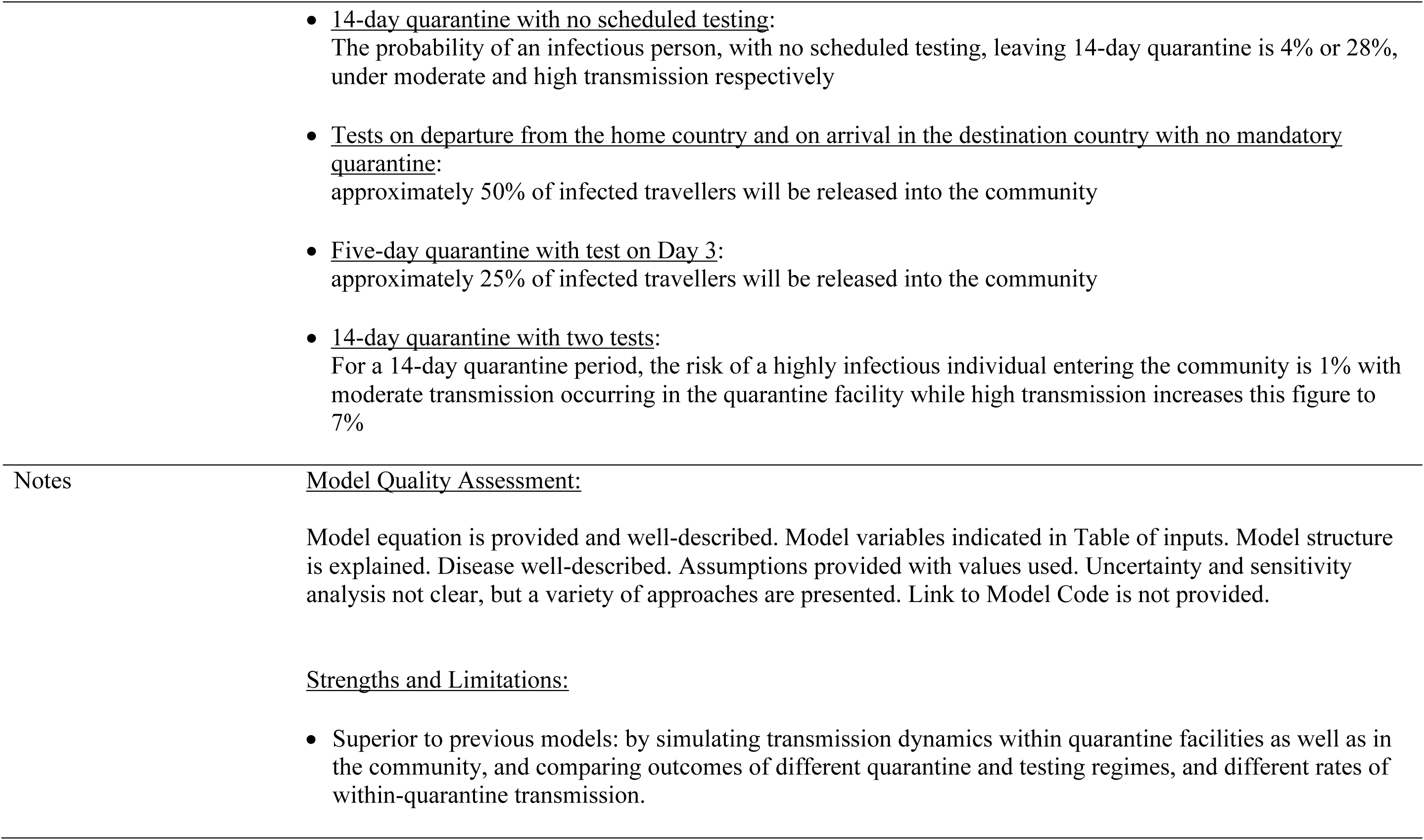
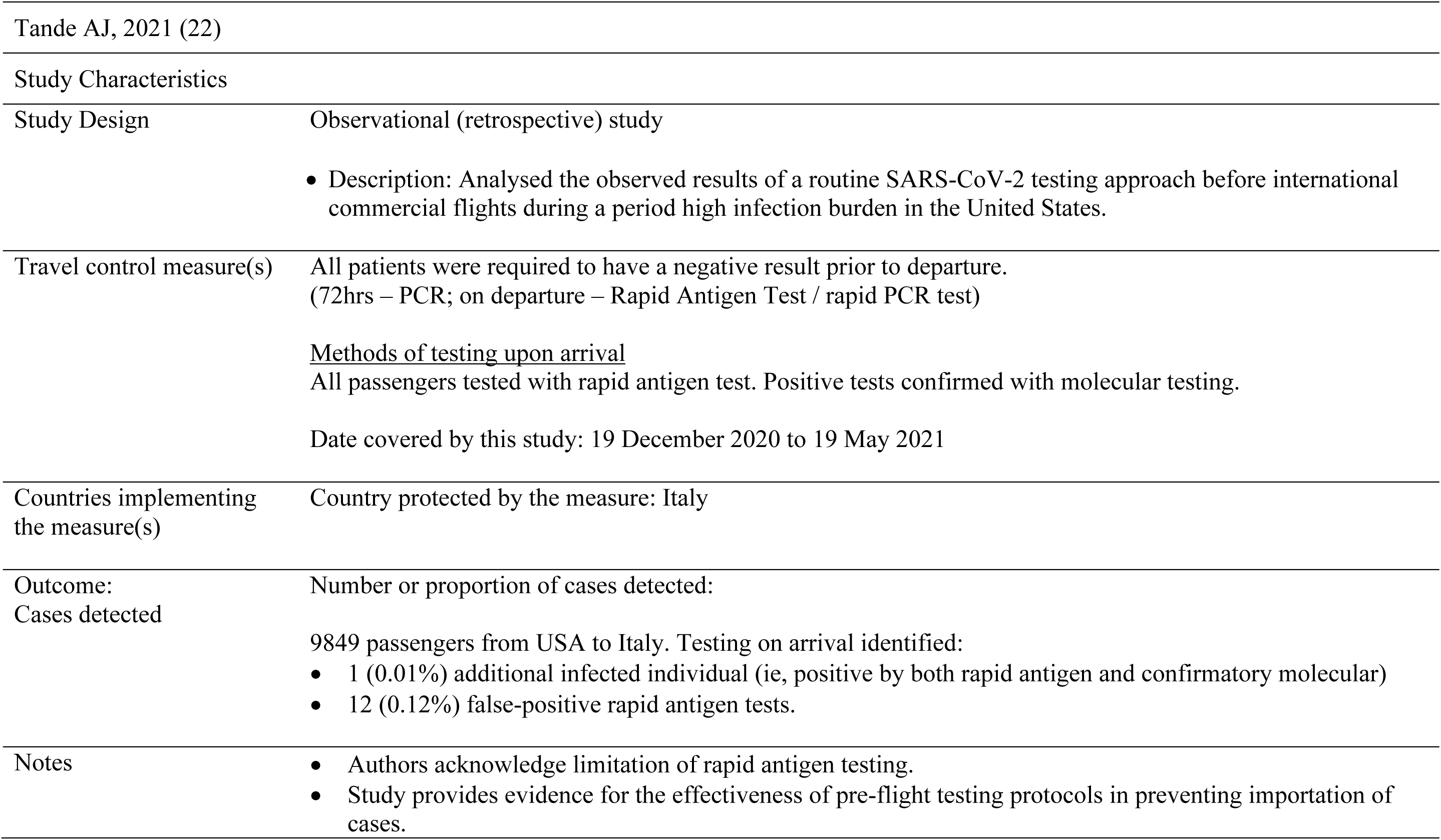
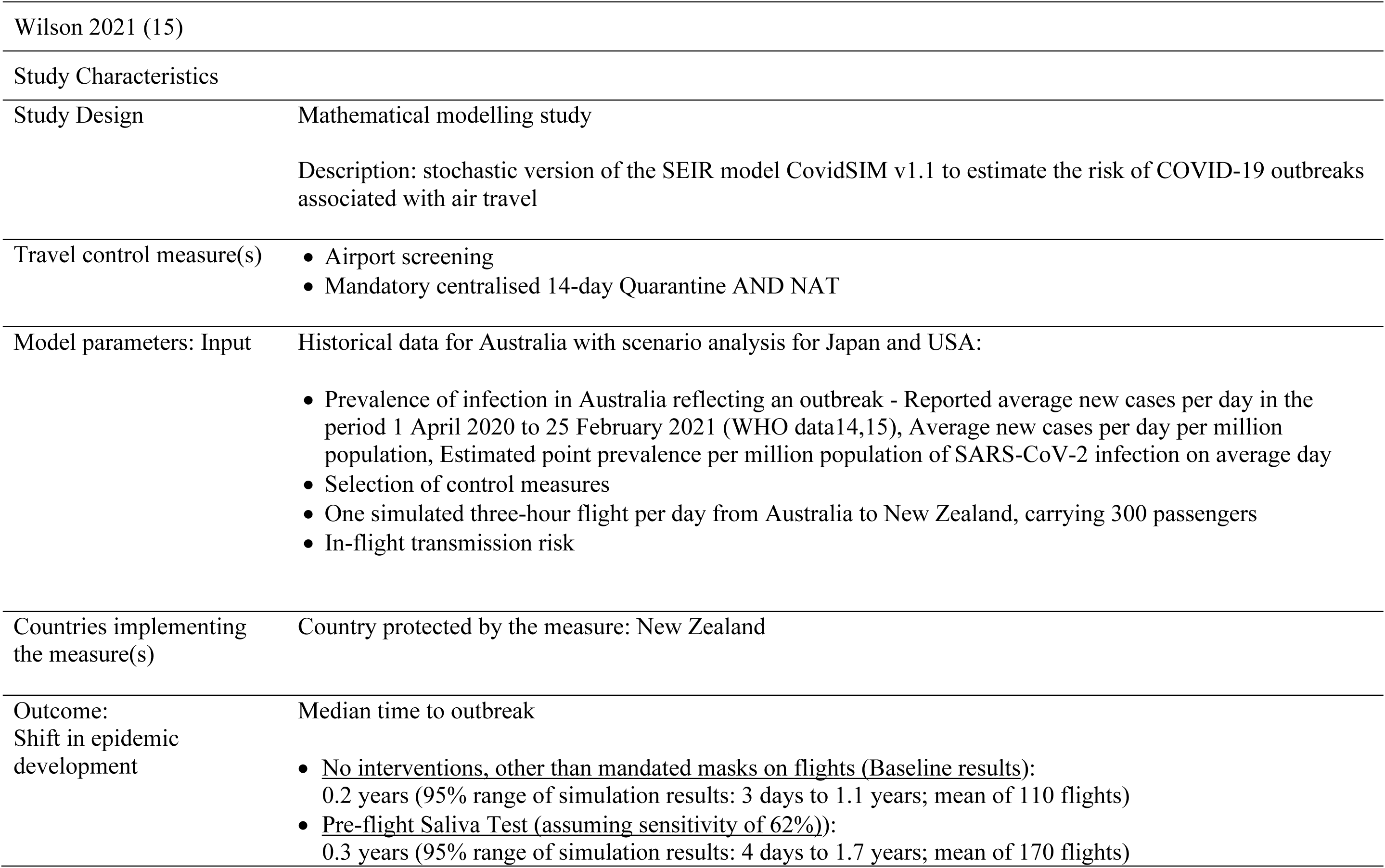
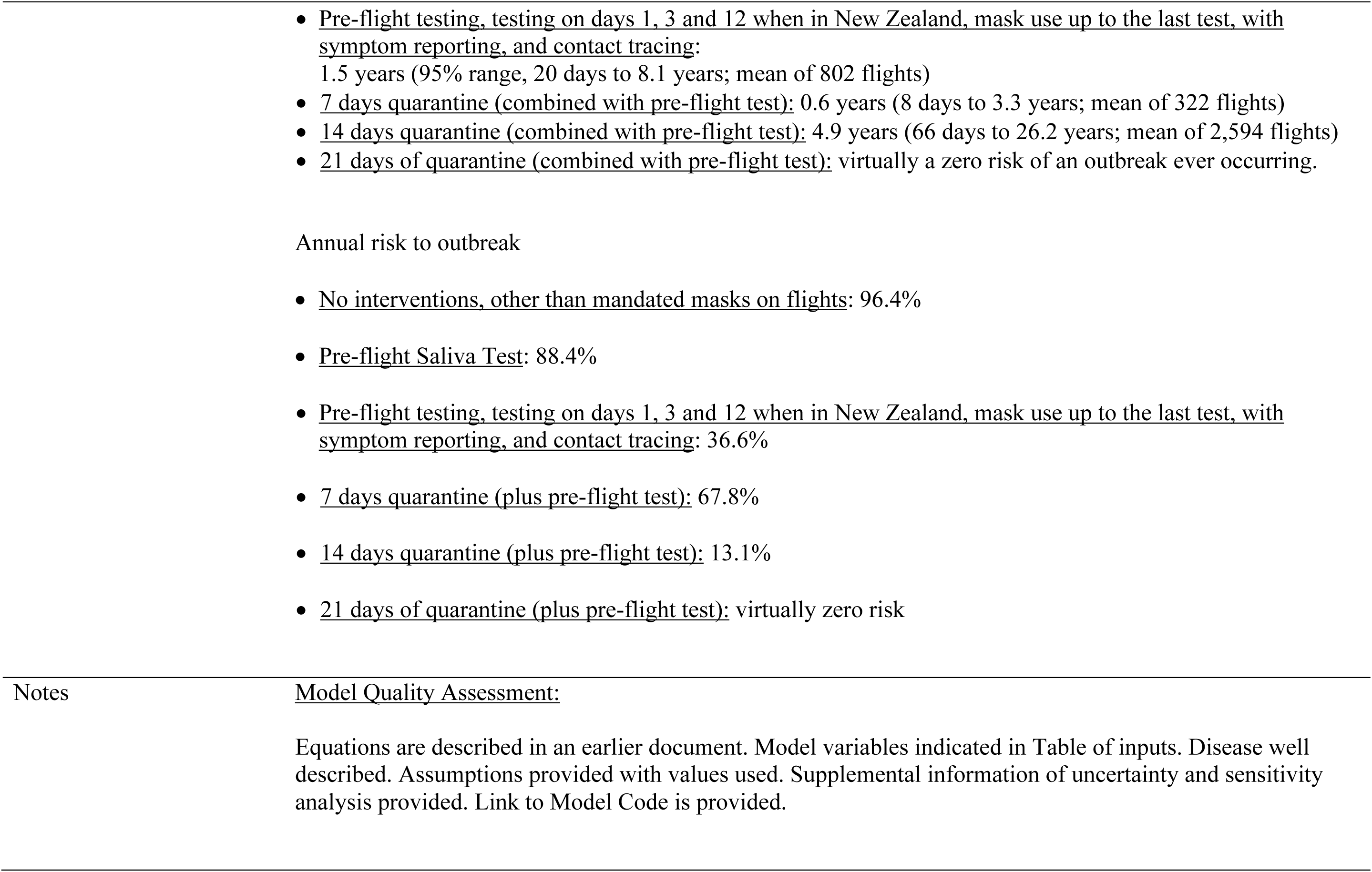
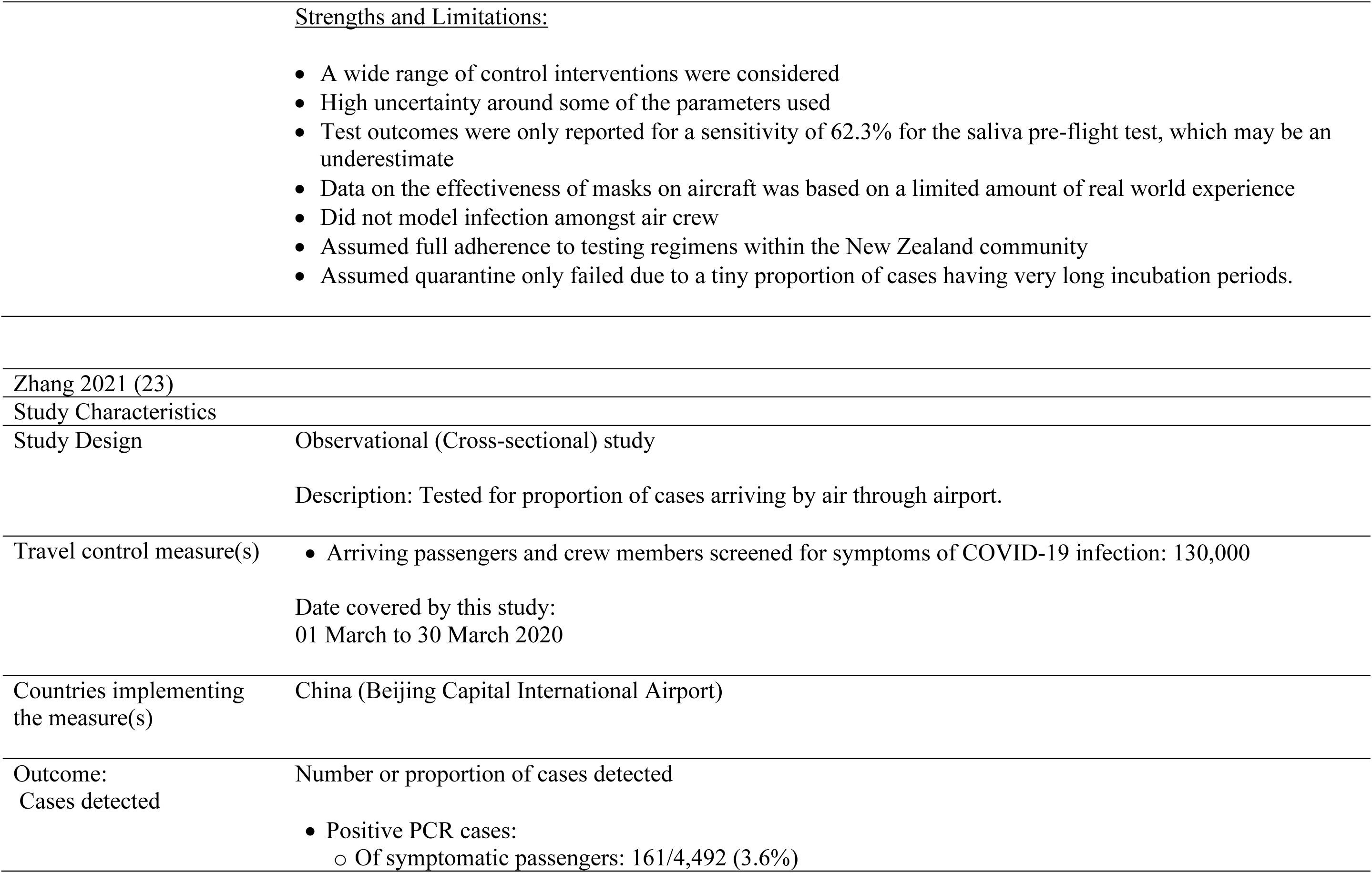
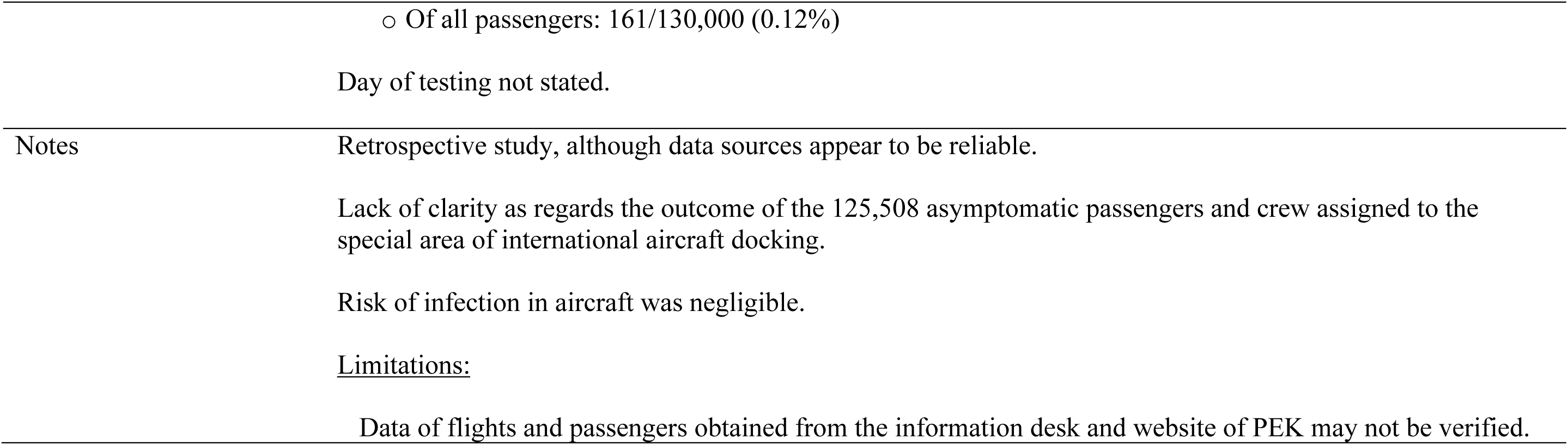
Characteristics of included studies [ordered by study ID]

**Table 5.**
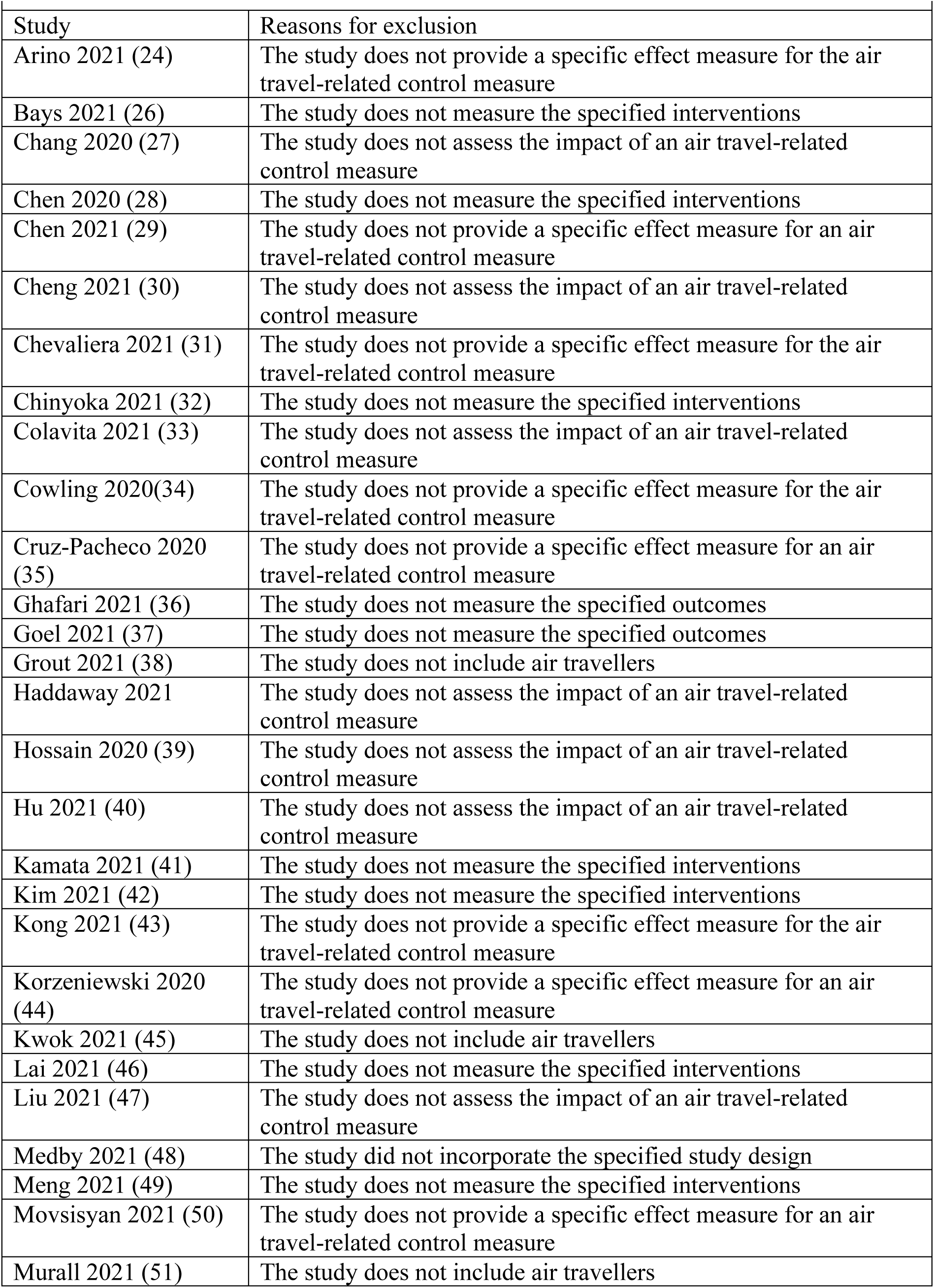
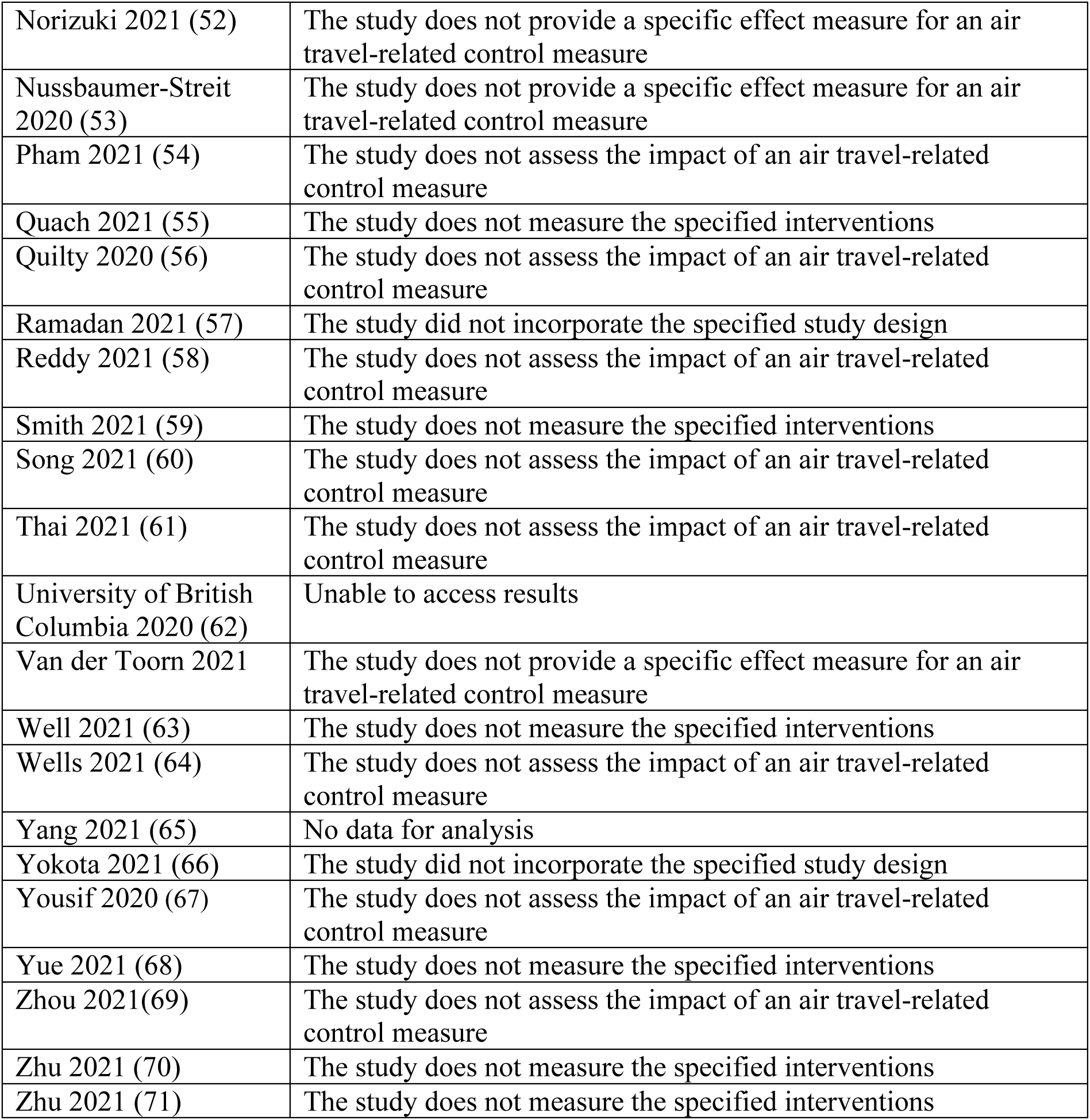
Characteristics of excluded studies [ordered by study ID]

### Study Designs

The updated search identified nine modelling studies (7–15) and eight observational studies (16–23).

### Risk of bias and quality of included studies

We summarised the included risk of bias for observational studies and quality of the modelling studies in Table 6 and Table 7, respectively. We used ROBINS-I to assess the risk of bias of observational studies concerned with microbiological testing, quarantine, and both. Overall, the risk of bias is deemed to be critical and moderate across five and three studies, respectively. Table 6 depicts the bias due to confounding, which was critical in three studies, moderate in one, with no information available in four. Bias in selection of participants into the study was moderate in five studies, critical in one, while no information was provided in the two remaining studies. Bias in classification of interventions was critical in three studies, low and moderate in one each, with no information provided in three studies. Bias due to deviations from intended interventions was critical in two studies, moderate in two, low in one, with no information in two. Bias due to missing data was critical in two studies, moderate in four studies, low in one and no information in three. Bias in measurement of outcomes was critical in one study, moderate in three, low in one and no information in three studies. Bias in selection of the reported result was critical in one study, low in four studies and no information in three studies. Amongst the modelling studies, three had validation concerns while one other failed to justify the structural assumptions.

**Table 6.** Summary of ROBINS-I risk of bias assessment for observational studies

| Domain | Abdulrahman<br>2021 | Cao-Lormeau<br>2021 | Fotheringham<br>2021 | O'Donnell<br>2021 | Ohlsen<br>2021 | Lunney<br>2021 | Tande<br>2021 | Zhang<br>2021 |
| --- | --- | --- | --- | --- | --- | --- | --- | --- |
| Bias due to confounding | Critical | Critical | NI | Critical | NI | NI | NI | Moderate |
| Bias in selection of participants into the study (external) | Moderate | Moderate | Moderate | Critical | NI | Moderate | NI | Moderate |
| Bias in classification of interventions | Critical | NI | NI | Critical | NI | Critical | Low | Moderate |
| Bias due to deviations from intended interventions | Critical | NI | NI | Critical | Moderate | NI | Low | Moderate |
| Bias due to missing data | Moderate | Critical | Critical | Moderate | NI | Moderate | Low | Moderate |
| Bias in measurement of outcomes | NI | NI | NI | Critical | Moderate | Moderate | Low | Moderate |
| Bias in selection of the reported result | Low | NI | NI | Critical | NI | Low | Low | Low |
| Overall RoB | Critical | Critical | Critical | Critical | Moderate | Critical | Moderate | Moderate |

**Table 7.**
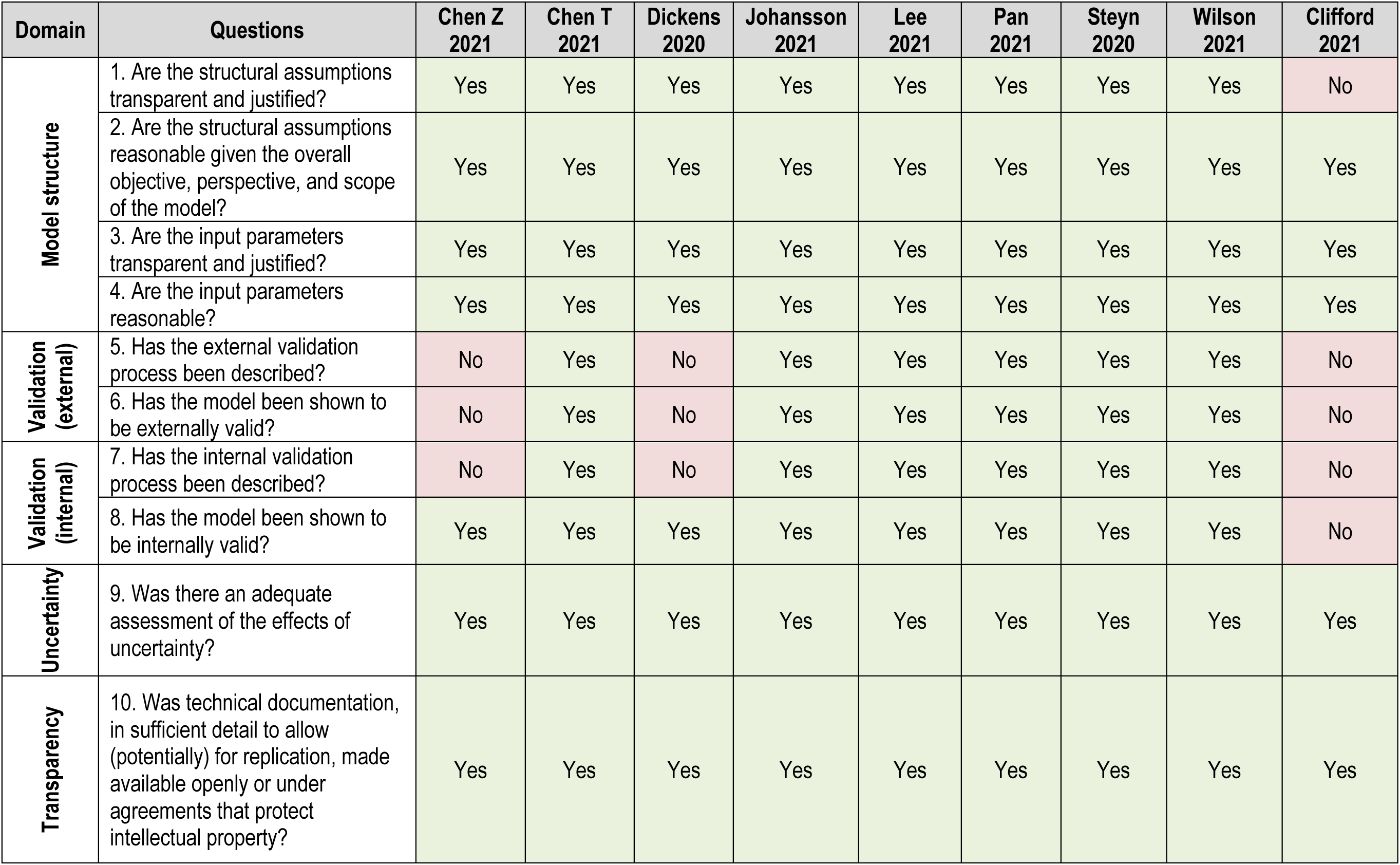
Summary of quality appraisal for modelling studies

### Outcomes

#### 1. Cases avoided due to the measure

##### Number or proportion of imported or exported cases

Together with three new modelling studies identified in the updated search (10, 12, 13), a total of four studies confirms that quarantine (with or without symptoms) and with no testing is effective in reducing the risk of transmission. Reduction in the proportion of imported cases ranged from 55% (7-days’ quarantine) to 99% (21 days’ quarantine) (very low certainty of evidence).

Two modelling studies on microbiological detection (10, 14) were identified in the updated search. Assuming that exposure had occurred, pre-departure testing was predicted to improve the detection of cases by up to 84.4%, although results vary according to timing of the test.

Two modelling studies (9, 10) demonstrated that a combination of quarantine and microbiological detection with a PCR test at day 7 significantly increases the proportion of cases averted as compared with quarantine alone, thus reducing the quarantine period.

#### 2. Shift in epidemic development

This outcome is subclassified into:

a. Time to diagnosis
b. Median time to outbreak
c. Risk of an outbreak

##### a. Time to diagnosis

One new modelling study (8) on quarantine alone and in combination with microbiological detection assesses time to diagnosis. The results suggest that these interventions would decrease the mean interval of diagnosis from arrival to laboratory-confirmation by up to 1.7 days. However, magnitude of effect may depend on testing sufficiency and lab capacity and policy implementation.

##### b. Median time to outbreak

One new modelling study (15) was identified in the updated search and reported that microbiological detection alone would delay an outbreak by 0.3 years. In combination with a 7-day quarantine or 21-day quarantine, the median time to outbreak respectively goes from to 0.6 years to virtual disappearance (very low-certainty evidence).

##### c. Risk of an outbreak

One new modelling study (15) reported that pre-flight testing together with mask usage, symptom reporting and contact tracing reduced the risk of an outbreak from 88% to 37% (very low-certainty evidence).

#### 3. Cases detected due to the measure

This outcome is subclassified into:

a. Proportion of cases detected
b. Reduction in local transmission
c. Probability of releasing an infected individual into the community

##### a. Proportion of cases detected

For this outcome, two observational studies on quarantine (19, 20), were identified in the updated search, thus adding to the earlier modelling study (24) which indicated an increasing positivity correlating with increased quarantine. Amongst participants released from quarantine following a negative test result on arrival, positive cases constituted in excess of 30% in both observational studies (very low-certainty evidence). However, generalisability of these results may be impacted given that passengers in these studies were not stratified according to air vs travel, and the second study comprised military-related conditions.

Five observational studies assessing the impact of microbiological detection only on this outcome (17, 19, 21–23) reported detecting from 0.01% to 1.5% of cases amongst all travellers. Self-collection of samples was conducted at 4 days post-arrival (17).

Two observational studies assessed the impact of a combination of quarantine and microbiological detection on this outcome, reporting positivity rates of 0.2% and 0.6% following 10-14 days of quarantine amongst >25,000 PCR-negative arrivals to Bahrain(16). The second study observed 26% positivity, with the majority detected within the first week (18).

##### b. Reduction in local transmission

Two modelling studies assessing the impact of isolation and quarantine on reducing local transmission (7, 11) were identified by the updated search. Implementing quarantine for all inbound travellers is the most efficient measure in reducing local cases by up to 23.3%. An even greater reduction (30 - 35%) was observed through immediate isolation following symptom onset prior to, or during, travel while quarantine implemented following exposure without symptom monitoring or testing, leads to a risk reduction from 64 - 95% (@7 days) to 96–100% (@14 days)

A single study (11) reported that pre-departure testing can reduce risk of transmission during travel by between 10 - 72% depending on time interval between testing and day of travel. Time of specimen collection, and degree of sensitivity of the test may introduce variability in the results.

Combination of interventions also had a single study (11) showing that on day 5–6: risk reduction = 97-100%.

##### c. Probability of releasing an infected individual into the community

No new studies were identified in the updated search. The findings of the earlier review of three modelling studies (25) (9) (14) reported quarantine as having a positive effect on the probability of releasing an infected individual into the community. Steyn indicated that the probability of effect varies according to moderate and high transmission conditions.

Combination of interventions had a single study (14) indicating that quarantine with testing of all incoming travellers would result in a risk of imported cases being released into the community of 25% following a 7-day quarantine period.

## Conclusions

We identified 17 studies which consisted modelling and observational studies in addition to the Cochrane review’s 31 studies. These studies compared the effectiveness of travel-related control measures (specifically quarantine, microbiological detection or a combination thereof) as a means to limit the spread of COVID-19. These additional studies do not alter the findings, nor the quality of the evidence (very low certainty) as found in the Cochrane Review. Due to the nature of GRADEing the certainty of the evidence for modelling and the type observational studies, it is unlikely that more studies with these designs will change the quality of the evidence. Generally, the data, with very low certainty of evidence, indicate that these interventions are effective. However, there is a need for higher quality observational and experimental studies to support our findings.

## Data Availability

All data produced in the present work are contained in the manuscript.

## Notes

### Competing Interest Statement

The authors have declared no competing interest.

### Funding Statement

This study was funded by World Health Organization and SPOR Evidence Alliance

